# Improving the quality of in-patient neonatal routine data as a pre-requisite for monitoring and improving quality of care at scale: A multi-site retrospective cohort study in Kenyan hospitals

**DOI:** 10.1101/2022.05.31.22275848

**Authors:** Timothy Tuti, Jalemba Aluvaala, Daisy Chelangat, George Mbevi, John Wainaina, Livingstone Mumelo, Kefa Wairoto, Dolphine Mochache, Grace Irimu, Michuki Maina, Mike English, The Clinical Information Network Group

## Abstract

**Objectives:** The objectives of this study were to (1) determine if membership of a clinical information network (CIN) was associated with an improvement in the quality of documentation of in-patient neonatal care provided over time, and (2) characterise accuracy of prescribing for basic treatments provided to neonatal in-patients if data are adequate.

**Design and Settings:** This was a retrospective cohort study involving all children aged ≤28 days admitted to New-Born Units (NBUs) between January 2018 and December 2021 in 20 government hospitals with an interquartile range of annual NBU inpatient admissions between 550 and 1640 in Kenya. These hospitals participated in routine audit and feedback processes on quality of documentation and care over the study period.

**Outcomes:** The study’s outcomes were the number of patients as a proportion of all eligible patients with (1) complete domain-specific documentation scores, and (2) accurate domain-specific treatment prescription scores at admission.

**Findings:** 80060 NBU admissions were eligible for inclusion. Upon joining the CIN, documentation scores in the *monitoring (vital signs)*, *other physical examination and bedside testing*, *discharge information*, and *maternal history* domains demonstrated a statistically significant month-to-month relative improvement in number of patients with complete documentation of 7.6%, 2.9%, 2.4%, and 2.0% respectively. There was also statistically significant month-to-month improvement in prescribing accuracy after joining the CIN of 2.8% and 1.4% for feeds and fluids but not for Antibiotic prescriptions. Findings suggest that much of the variation observed is due to hospital-level factors.

**Conclusions:** It is possible to introduce tools that capture important clinical data at least 80% of the time in routine African hospital settings but analyses of such data will need to account for missingness using appropriate statistical techniques. These data allow trends in performance to be explored and could support better impact evaluation, performance benchmarking, exploration of links between health system inputs and outcomes and scrutiny of variation in quality and outcomes of hospital care.

## Introduction

Neonatal (new-born children aged ≤ 28 days) deaths account for 47% of all under-five deaths with 37% of these deaths occurring in Sub-Saharan African (SSA) countries [1]. These deaths are largely attributable to preterm birth, sepsis and intrapartum complications[2] and hospital admissions with these conditions are still associated with high morbidity in Low- and Middle-Income Countries (LIMCs) like Kenya [3]. Essential interventions such as newborn resuscitation, Kangaroo Mother Care (KMC), early recognition and treatment of neonatal infections, and Continuous Positive Airway Pressure (CPAP) therapy have been identified as major interventions to reduce neonatal deaths in hospitals [4, 5]. However, available evidence suggests that adherence to recommended care giving practices in LMICs is poor [6–8] while poorly functioning information systems mean limited data of questionable quality on the delivery of such interventions in routine hospital settings in LMIC is available [9, 10].

Availability of high-quality timely, accessible, and easy to use data from routine clinical settings could improve monitoring of intervention adoption and quality of hospital care at scale, and ultimately might help improve clinical outcomes [9–12]. An integrated approach providing a mechanism to promote continued improvement of clinical information, implementation of effective practices and technologies, and locally relevant research can comprise a ‘learning health system’, which are posited to be influential in producing the positive change required [13–16].

The objectives of this study were to determine: (1) if good quality data can be generated from hospitals’ newborn units invited to participate in a low-cost learning health system and whether the quality of documentation that is the source of routine data improves over time, (2) if basic recommended treatments or interventions are being correctly provided to neonatal in-patients (if the quality of clinical data permits this) and (3) the potential of data to support tracking of intervention adoption and ultimately their effects in LMIC.

## Methods

### Ethics and Reporting

The reporting of this observational study follows the Strengthening of reporting of observational studies in epidemiology (STROBE) statement [17]. The Scientific and Ethics Review Unit of the Kenya Medical Research Institute (KEMRI) approved the collection of the de-identified data that provides the basis for this study as part of the Clinical Information Network (CIN) for newborns (CIN-N). Individual consent for access to de-identified patient data was not required.

### Study design and setting

This study is situated within the CIN-N. The CIN-N is a collaborative learning health system network between KEMRI-Wellcome Trust Research Programme, the Ministry of Health, Kenya Paediatric Association, and 21 partner hospitals [9, 18]. The hospitals in CIN-N are first referral-level, geographically dispersed hospitals with an interquartile range of annual NBU inpatient admissions between 550 and 1640. A paediatric network was established in 2013/2014 to improve care given to inpatient children [13]. After co-development work with a single large NBU, multiple hospitals’ neonatal units joined to extend the original paediatric network and create the CIN-N in 2017/2018. In these hospitals most admission care and prescribing is done by medical officer interns who rotate departments regularly resulting in almost complete changes in those responsible for NBU admissions every three months [19]. In-depth description of the development of CIN-N is detailed elsewhere [3, 9, 18]. For the purposes of this study, the hospital which was instrumental in the design of the data collection tools for better capture of NBU data is omitted from subsequent analyses because it was developing and using information tools for four years before any additional hospitals joined the CIN-N; thus, only data from 20 hospitals is analysed.

This was a retrospective cohort study involving NBUs in the CIN-N hospitals. The CIN-N hospitals receive three-monthly clinical audit and feedback reports on the quality of care including for example, summaries of key issues for documentation and treatment prescription errors once they join [18]. Shorter feedback reports on data quality, and morbidity and mortality reports are disseminated monthly via email to clinicians, nurses in-charge and other hospital administration staff. Neonatal team leaders (neonatologists, paediatricians, and nurses) met face to face once or twice annually until 2020 (before the COVID-19 pandemic) to discuss these reports and how to improve clinical care. Finally, those that received no feedback were neither included in written reports nor discussed in meetings. During the COVID-19 pandemic only short, online network meetings were conducted that focused mostly on disseminating information of relevance to the pandemic and efforts to improve local neonatal audit and nursing practices.

### Study size and participants

All hospitals have a specific newborn unit (NBU) and all aged ≤28 days admitted (1) between January 2018 and December 2021 to the NBU of 20 CIN-N hospitals were eligible for inclusion. We excluded neonates whose admission or discharge dates were missing or improbable (e.g. discharge date is earlier than admission date), and those whose admissions fell within prolonged health worker strikes that resulted in major disruption to health care delivery (i.e. December 2020 – January 2021) [20].

### Data sources and management

Methods of collection and cleaning of data in the CIN-N are reported in detail elsewhere [21]. Clinical data for neonatal admissions to the hospitals within the CIN-N are captured through Neonatal Admission Record (NAR) forms and other forms and charts that are part of the hospital’s medical record. The NAR and associated patient charts prompt the clinician with a checklist of fields covering nine documentation domains that include demographics, admission information, discharge information, maternal history, presenting complaints, cardinal signs on examination, other physical examinations, nursing monitoring and supportive care [18]. Other charts that are also used are a comprehensive newborn monitoring chart (collects data on vital signs, feeds and fluids prescribed) which were developed and introduced between March and June 2019 [22], transfer forms (containing key data when a baby is transferred internally from maternity unit to NBU), treatment sheets, discharge summaries and death notification reports in case death occurs.

The clinical signs included in the NAR are based on recommendations in guidelines from the national Ministry of Health and the World Health Organisation (WHO) [23]. NAR forms were originally developed as part of the Emergency Treatment and Triage plus admission (ETAT+) approach which includes skill training in essential inpatient newborn care [24]. In earlier work they were associated with improved documentation of key patient characteristics during admission [18]. NAR are not provided to hospitals in CIN-N and so their adoption is at the discretion of hospital teams and supported by hospitals’ own resources, although CIN-N hospitals are encouraged to use them.

Each hospital has a clerk who extracts data from the NAR forms into a Research Electronic Data Capture (REDCap) database [25]. Two sets of data are captured: minimum and full datasets. The minimal dataset – which is unsuitable for this study’s analyses - is collected for (1) admissions during major holidays when the data clerk is on leave, and (2) on a random selection of records in hospitals where the workload is very high. The minimal dataset includes biodata and patient outcomes at discharge and is collected on all neonatal admissions in all CIN-N hospitals for reporting to the national Health Information System. The full dataset contains comprehensive data on admission details, patient history, clinical investigations, treatment and discharge information including diagnoses and outcome [9]. The data collected is subjected to routine quality assurance checks explained in detail elsewhere [9].

### Quantitative variables

#### Creation of documentation scores

The outcome for objective 1 of this study was based on use of individual patient documentation scores compiled from the signs, symptoms, treatments, and outcomes data (Table 1). These scores were developed for each of the eight NAR indicator documentation domains then used to determine trends in the completeness of documentation in the hospitals involved. Domains had different numbers of component data items (Table 1) considered key for characterising NBU populations and assessing core aspects of technical quality of care neonates receive [26]. Domain-specific composite scores for each patient were developed by arithmetic aggregation of all items with valid (non-missing) data in that domain (score = 1 if valid data, = 0 for missing data).

**Table 1:**
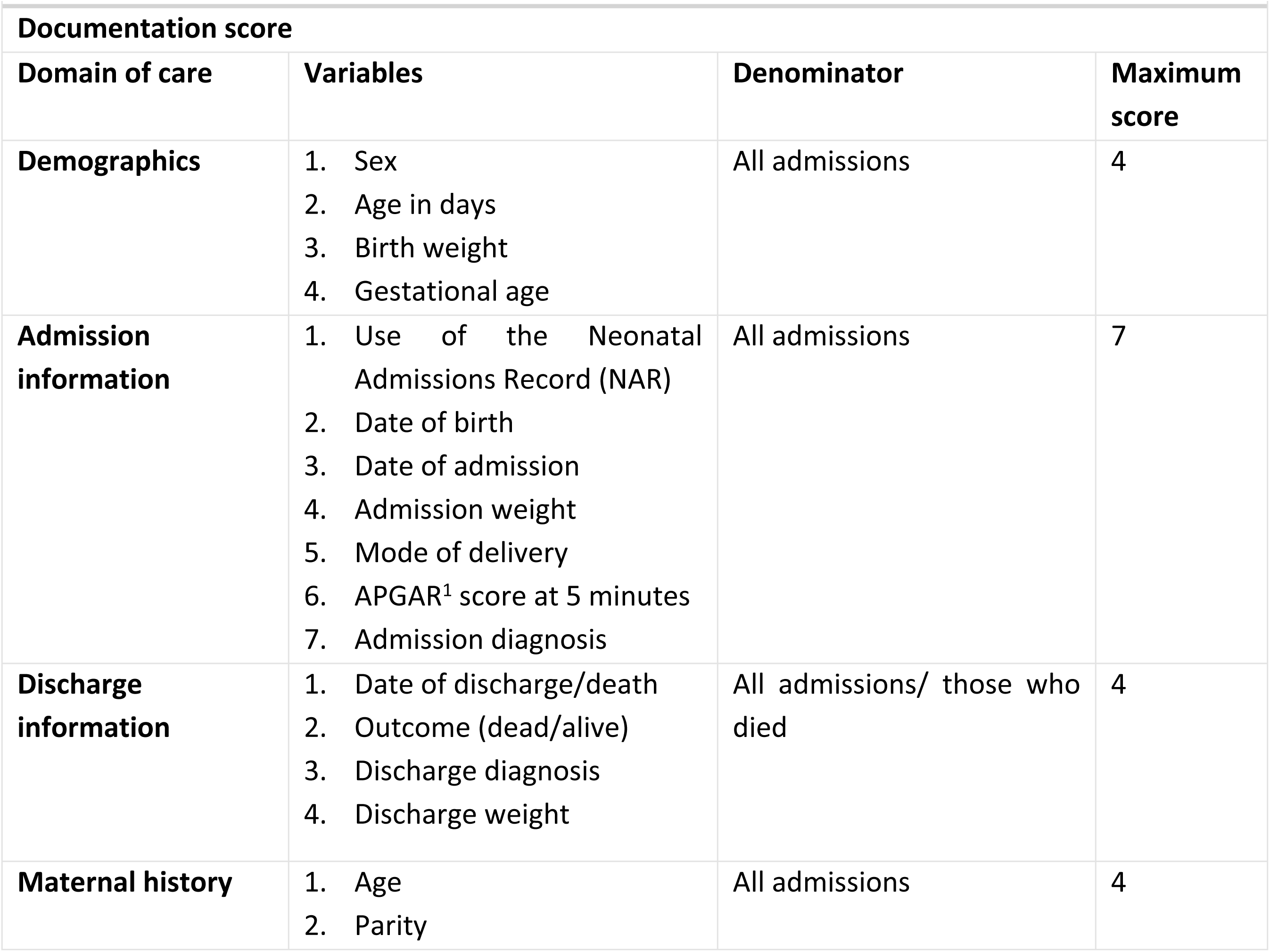
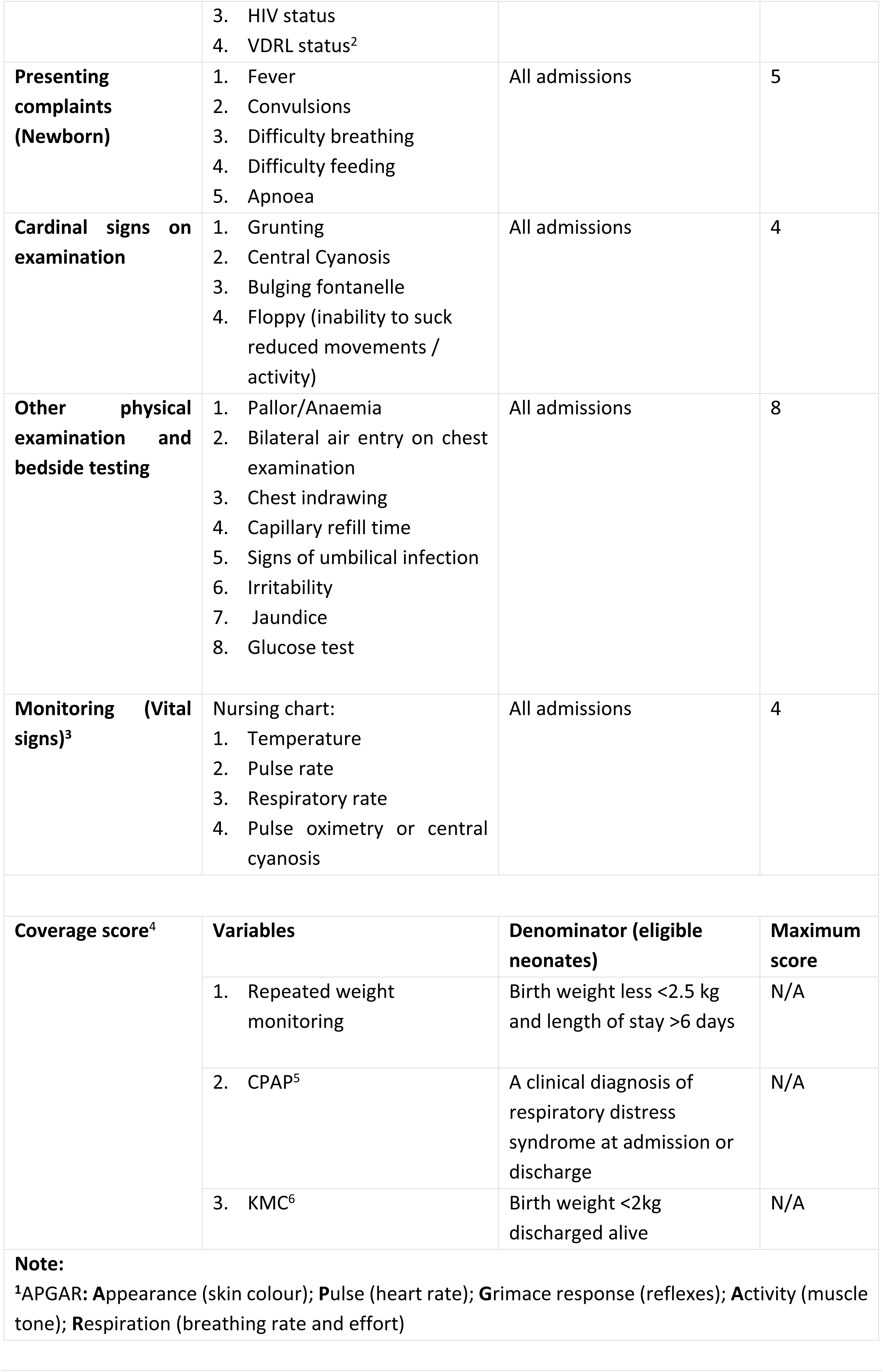
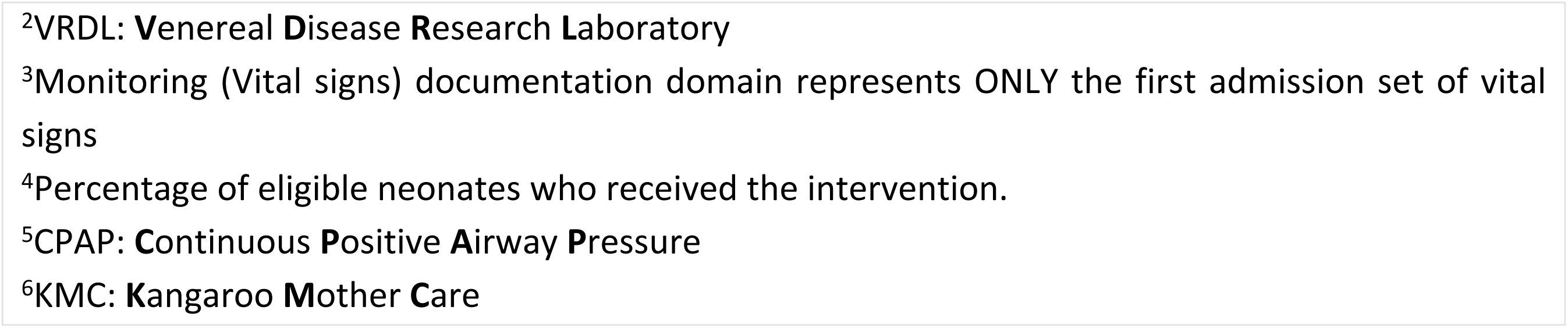
Variables used in the documentation and coverage scores

| **Documentation score** |  |  |  |
| --- | --- | --- | --- |
| **Domain of care** | **Variables** | **Denominator** | **Maximum score** |
| **Demographics** | Sex<br>Age in days<br>Birth weight<br>Gestational age | All admissions | 4 |
| **Admission information** | Use of the Neonatal Admissions Record (NAR)<br>Date of birth<br>Date of admission<br>Admission weight<br>Mode of delivery<br>APGAR<sup>1</sup> score at 5 minutes<br>Admission diagnosis | All admissions | 7 |
| **Discharge information** | Date of discharge/death<br>Outcome (dead/alive)<br>Discharge diagnosis<br>Discharge weight | All admissions/ those who died | 4 |
| **Maternal history** | Age<br>Parity | All admissions | 4 |
|  | 3. HIV status<br>4. VDRL status<sup>2</sup> |  |  |
| **Presenting complaints (Newborn)** | 1. Fever<br>2. Convulsions<br>3. Difficulty breathing<br>4. Difficulty feeding<br>5. Apnoea | All admissions | 5 |
| **Cardinal signs on examination** | 1. Grunting<br>2. Central Cyanosis<br>3. Bulging fontanelle<br>4. Floppy (inability to suck reduced movements / activity) | All admissions | 4 |
| **Other physical examination and bedside testing** | 1. Pallor/Anaemia<br>2. Bilateral air entry on chest examination<br>3. Chest indrawing<br>4. Capillary refill time<br>5. Signs of umbilical infection<br>6. Irritability<br>7. Jaundice<br>8. Glucose test | All admissions | 8 |
| **Monitoring (Vital signs)<sup>3</sup>** | Nursing chart:<br>1. Temperature<br>2. Pulse rate<br>3. Respiratory rate<br>4. Pulse oximetry or central cyanosis | All admissions | 4 |
| **Coverage score<sup>4</sup>** | **Variables** | **Denominator (eligible neonates)** | **Maximum score** |
|  | 1. Repeated weight monitoring | Birth weight less <2.5 kg and length of stay >6 days | N/A |
|  | 2. CPAP<sup>5</sup> | A clinical diagnosis of respiratory distress syndrome at admission or discharge | N/A |
|  | 3. KMC<sup>6</sup> | Birth weight <2kg discharged alive | N/A |
| **Note:**<br><sup>1</sup>APGAR: Appearance (skin colour); Pulse (heart rate); Grimace response (reflexes); Activity (muscle tone); Respiration (breathing rate and effort) |  |  |  |

<sup>2</sup>VRDL: **V**enereal **D**isease **R**esearch **L**aboratory
<sup>3</sup>Monitoring (Vital signs) documentation domain represents ONLY the first admission set of vital signs
<sup>4</sup>Percentage of eligible neonates who received the intervention.
<sup>5</sup>CPAP: **C**ontinuous **P**ositive **A**irway **P**ressure
<sup>6</sup>KMC: **K**angaroo **M**other **C**are

#### Creation of treatment correctness scores and intervention tracking

We created an additional three indicator domains to reflect the accuracy of basic treatment prescriptions (antibiotics, fluids, and feeds) for relevant sub-populations of neonates receiving these treatments and based on the dosage or volume recommendations in the national guidelines [27] (Table 2). Each eligible patient in each of the treatment domains in the analysis could either have correct or incorrect prescription: If the treatment was correctly prescribed, then it contributed a score of one to the domain-specific score; if treatment information was missing or treatment was incorrectly prescribed, then it contributed a score of zero to the domain-specific score. Finally, we descriptively assessed how well the data could support tracking of intervention adoption over time by evaluating whether neonates eligible for weight monitoring, CPAP, and KMC received these essential services. The adoption of weight monitoring, CPAP and KMC were summarised by coverage scores calculated as the percentage of neonates potentially eligible who were recorded as receiving these interventions/monitoring.

**Table 2:**
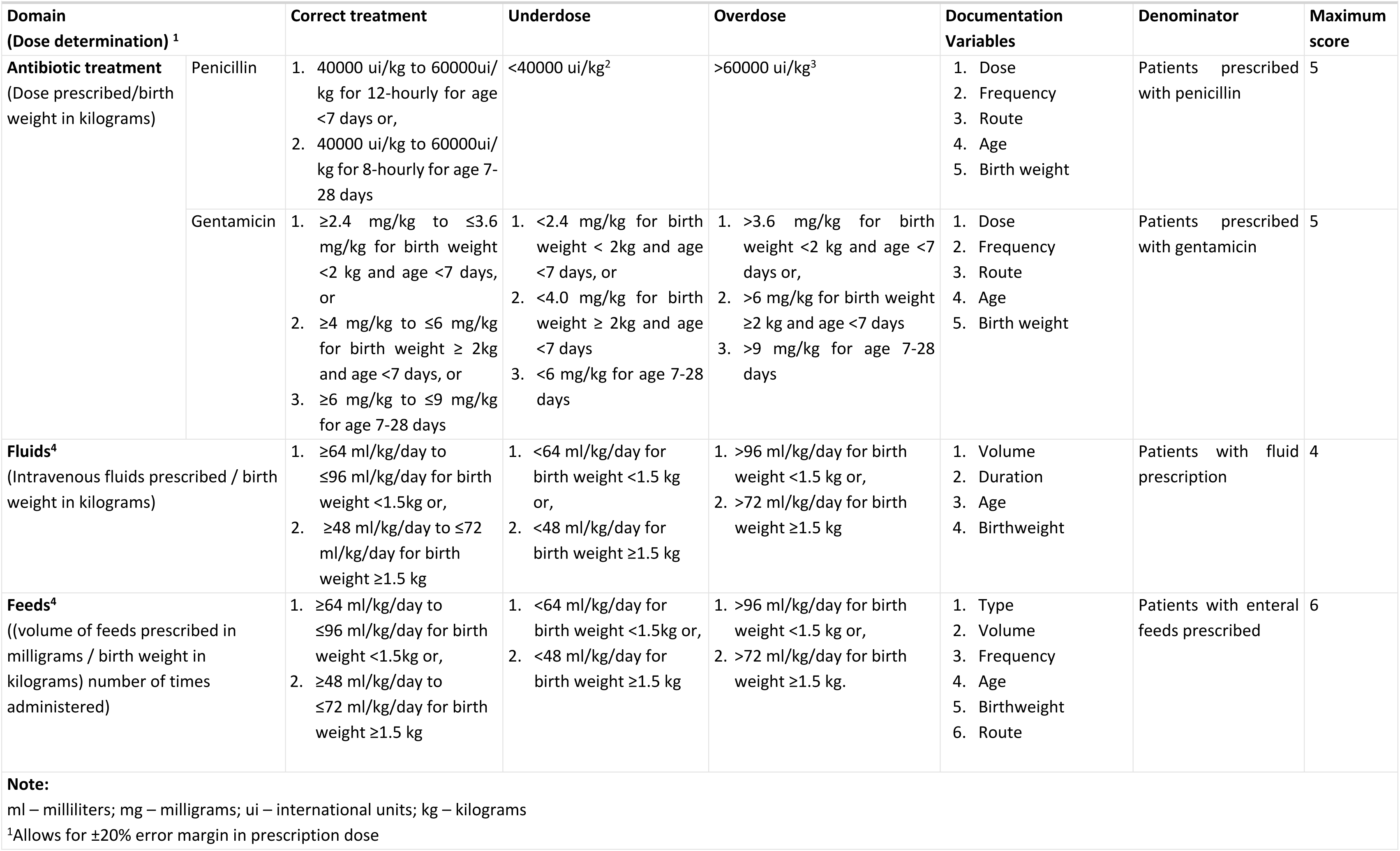
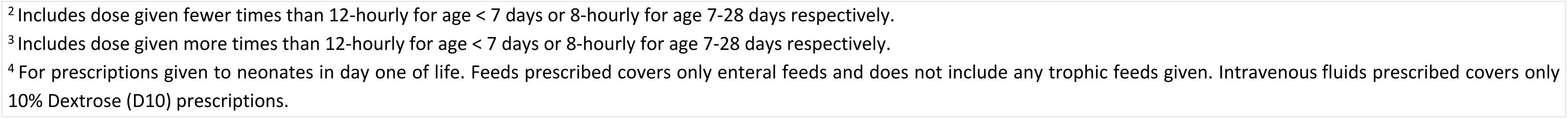
Threshold for prescribing antibiotics, fluids, and feeds

| Domain<br>(Dose determination) <sup>1</sup> |  | Correct treatment | Underdose | Overdose | Documentation Variables | Denominator | Maximum score |
| --- | --- | --- | --- | --- | --- | --- | --- |
| **Antibiotic treatment**<br>(Dose prescribed/birth weight in kilograms) | Penicillin | 1. 40000 ui/kg to 60000ui/kg for 12-hourly for age <7 days or,<br>2. 40000 ui/kg to 60000ui/kg for 8-hourly for age 7-28 days | <40000 ui/kg<sup>2</sup> | >60000 ui/kg<sup>3</sup> | 1. Dose<br>2. Frequency<br>3. Route<br>4. Age<br>5. Birth weight | Patients prescribed with penicillin | 5 |
|  | Gentamicin | 1. ≥2.4 mg/kg to ≤3.6 mg/kg for birth weight <2 kg and age <7 days, or<br>2. ≥4 mg/kg to ≤6 mg/kg for birth weight ≥ 2kg and age <7 days, or<br>3. ≥6 mg/kg to ≤9 mg/kg for age 7-28 days | 1. <2.4 mg/kg for birth weight < 2kg and age <7 days, or<br>2. <4.0 mg/kg for birth weight ≥ 2kg and age <7 days<br>3. <6 mg/kg for age 7-28 days | 1. >3.6 mg/kg for birth weight <2 kg and age <7 days or,<br>2. >6 mg/kg for birth weight ≥2 kg and age <7 days<br>3. >9 mg/kg for age 7-28 days | 1. Dose<br>2. Frequency<br>3. Route<br>4. Age<br>5. Birth weight | Patients prescribed with gentamicin | 5 |
| **Fluids<sup>4</sup>**<br>(Intravenous fluids prescribed / birth weight in kilograms) |  | 1. ≥64 ml/kg/day to ≤96 ml/kg/day for birth weight <1.5kg or,<br>2. ≥48 ml/kg/day to ≤72 ml/kg/day for birth weight ≥1.5 kg | 1. <64 ml/kg/day for birth weight <1.5 kg or,<br>2. <48 ml/kg/day for birth weight ≥1.5 kg | 1. >96 ml/kg/day for birth weight <1.5 kg or,<br>2. >72 ml/kg/day for birth weight ≥1.5 kg | 1. Volume<br>2. Duration<br>3. Age<br>4. Birthweight | Patients with fluid prescription | 4 |
| **Feeds<sup>4</sup>**<br>((volume of feeds prescribed in milligrams / birth weight in kilograms) number of times administered) |  | 1. ≥64 ml/kg/day to ≤96 ml/kg/day for birth weight <1.5kg or,<br>2. ≥48 ml/kg/day to ≤72 ml/kg/day for birth weight ≥1.5 kg | 1. <64 ml/kg/day for birth weight <1.5kg or,<br>2. <48 ml/kg/day for birth weight ≥1.5 kg | 1. >96 ml/kg/day for birth weight <1.5 kg or,<br>2. >72 ml/kg/day for birth weight ≥1.5 kg. | 1. Type<br>2. Volume<br>3. Frequency<br>4. Age<br>5. Birthweight<br>6. Route | Patients with enteral feeds prescribed | 6 |
| **Note:**<br>ml – milliliters; mg – milligrams; ui – international units; kg – kilograms<br><sup>1</sup>Allows for ±20% error margin in prescription dose |  |  |  |  |  |  |  |

<sup>2</sup> Includes dose given fewer times than 12-hourly for age < 7 days or 8-hourly for age 7-28 days respectively.
<sup>3</sup> Includes dose given more times than 12-hourly for age < 7 days or 8-hourly for age 7-28 days respectively.
<sup>4</sup> For prescriptions given to neonates in day one of life. Feeds prescribed covers only enteral feeds and does not include any trophic feeds given. Intravenous fluids prescribed covers only 10% Dextrose (D10) prescriptions.

### Statistical methods

Documentation performance of each hospital over time was summarised monthly using trend plots as a percentage representing (1) the average score of individual patient domain-specific score as a proportion of the maximum domain score possible (i.e., Documentation score), and (2) the number of patients with maximum possible documentation score for each domain out of all patients with full data collection admitted to the NBU. Similarly, domain-specific treatment accuracy and treatment coverage was summarised monthly for each hospital using trend plots as a percentage representing the proportion of neonates eligible for essential treatments who received an accurate prescription or required intervention respectively.

These pooled scores are presented using scatter plots for each hospital each month over the period 2018 to 2021 (i.e., from the month of joining CIN-N). We used Locally Weighted Scatterplot Smoothing (LOWESS) line plots to visually represent the trend over time for each documentation, treatment accuracy, and treatment coverage domain.

To characterise clinical documentation completeness and adherence to recommended treatment prescribing guidelines over time while quantifying heterogeneity between CIN-N hospitals, we fitted generalised linear mixed models for each documentation and treatment accuracy domain. These mixed models (using log link) were fitted on two types of hospital-level count outcome variables computed monthly:

(a) Documentation domain completeness score per hospital (as the number of all patients with all domain-specific variables documented out of all patients admitted to CIN-N hospitals).
(b) Treatment domain accuracy score per hospital (as the number of all patients with accurate treatment prescription out of all those with the treatment prescribed).

Our approach to the mixed models fitting is within the multilevel modelling framework, with patients nested in hospitals nested in time points. Time elapsed was captured as months since the hospitals joined the CIN-N and was treated as a continuous fixed effect. From previous studies within the CIN, the effect of time on adherence to recommended clinical practice was found to vary across hospitals [28]. We used a likelihood ratio test (LRT) to determine the most suitable random effects model (hospital random intercepts vs hospital random intercepts with random slopes for time). The outcome variables for documentation and treatment domains were assumed to follow a Poisson distribution. An offset term (i.e., the number or patients eligible per month per hospital) was included in each model to model the count outcome as a rate over time (e.g., change in the number of patients with accurate treatment prescribed), and the model effects reported as incidence rate ratios (IRR). We provide Intra-cluster correlation coefficients (ICC) to indicate variation between hospitals in recommended documentation practices and adherence to treatment guidelines.

### Missing data

Missing data was considered ‘informative’ as the analysis is based on documentation or no documentation. For the documentation score, missing variables were recorded as zero and therefore contributed a score of zero to the domain score per patient. For treatments, the absence of clear prescription information was logically considered to represent an inadequate prescription; for coverage, no record of use of the intervention was assumed to indicate no use.

### Sensitivity analyses

Overdispersion of the outcome variables (which is when the conditional variance exceeds the conditional mean), a key negative binomial model assumption, was evaluated by a likelihood ratio test comparing the model(s) to their Poisson model equivalent, which holds the conditional mean and variance to be equal (i.e., Equidispersion). Also, a likelihood ratio test (LRT) was used to examine the most suitable random effect model (random intercepts at the hospital level versus random intercepts for the hospitals with random slopes for time) [29]. To ensure that we adequately reflect the correlations between the repeated outcome measurements of each hospital which decrease with time lag (i.e. autocorrelation), we used LRT to examine if there would be evidence to support including a term for an autocorrelation structure of order one [30], over using a mixed model without such a term.

Finally, we explored whether there was evidence supporting the assumption that the conditional outcome of the models’ approximated a normal distribution using quantile residual quantile-quantile (QQ) plot for each fitted model, although this assumption is debatable for count data models [31].

## Results

### Descriptive findings

Figure 1 depicts the study population inclusion process. Out of the 84,960 NBU admissions to CIN-N hospitals, 80,060 (94.23%) were eligible for analysis. Most exclusions were because an admission was randomly sampled for minimum data collection (2934/84960) or fell in industrial action period (1966/84960). Among the patients admitted to CIN-N during the study period and selected for this study, 43953/80060 (54.9%) were male. Overall, the mortality rate across the 20 hospitals was 11314/80060 (14.13%). The median birth weight of CIN-N NBU admissions was 3 kgs (inter-quartile range (IQR): 2.0-3.395) and median length of stay was 4 days (1QR: 2-8). NBU admissions had a median of one admission diagnosis (IQR: 1-2). The leading NBU discharge diagnoses over time was low birth weight followed by birth asphyxia, respiratory distress syndrome, and then neonatal sepsis. Out of the 42998/80060 (53.71%) NBU admissions with Gentamicin prescription and 43889/80060 (54.82%) with Penicillin prescription, 1022/42998 (2.38%) and 1964/43889 (4.47%) were classified as incorrect because of incomplete prescribing data (e.g., any of missing age, birth weight, dosage, route, and frequency of administration variables) respectively. Out of the 35295/80060 (44.09%) NBU admissions with fluids prescription and 13643/80060 (17.04%) with feeds prescription, 2778/35295 (7.87%) and 3721/13643 (27.27%) were classified as incorrect because of incomplete prescribing data respectively. The proportion of records in which key items are not recorded is illustrated in Table 3.

**Figure 1:**
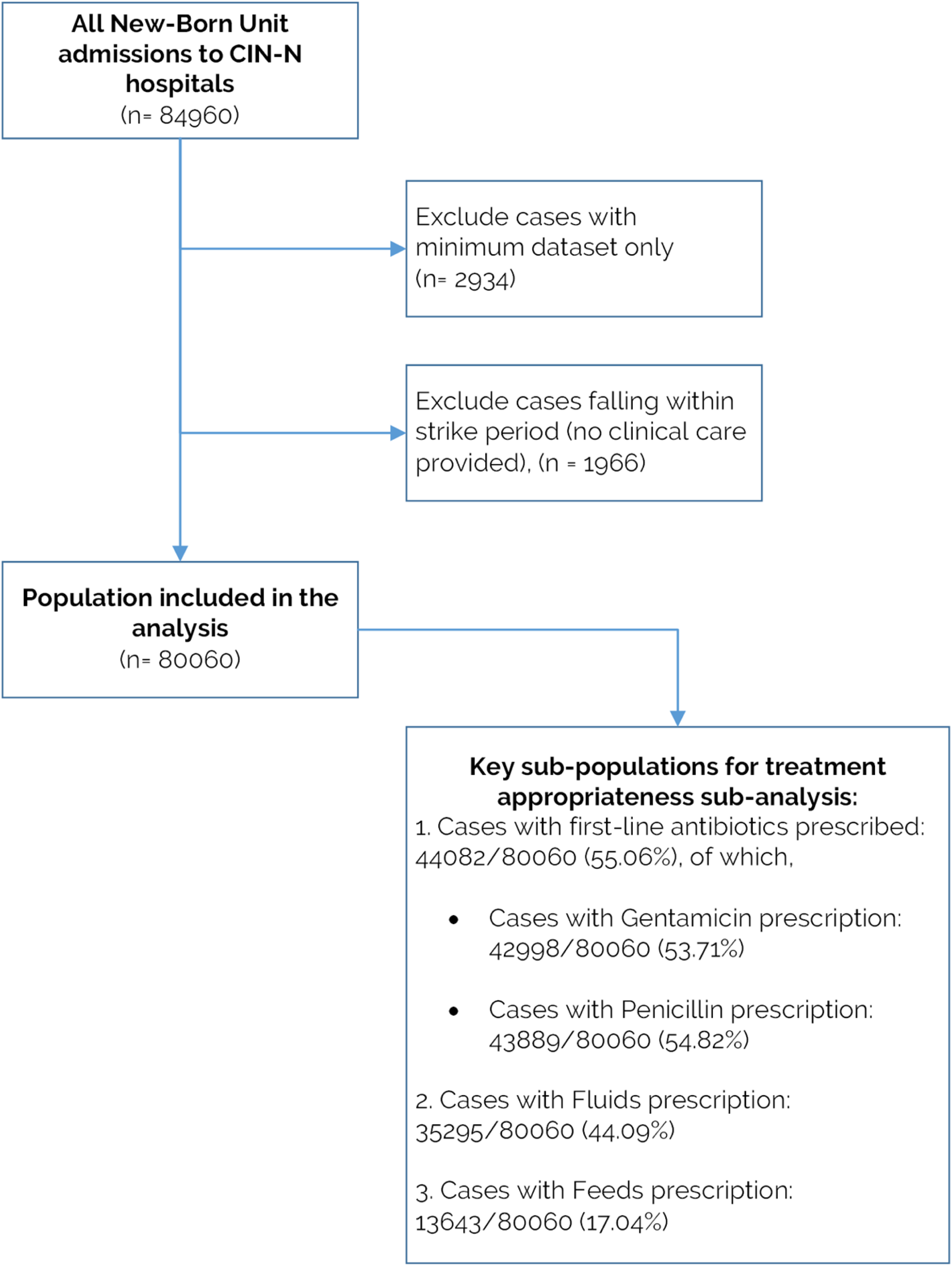
Flow-chart of the inclusion criteria. The overall population (n = 80060) is used for documentation score analysis. Supplementary Table 1 provides details of when CIN-N hospitals joined the network and patient records per hospital so far.

**Table 3:**
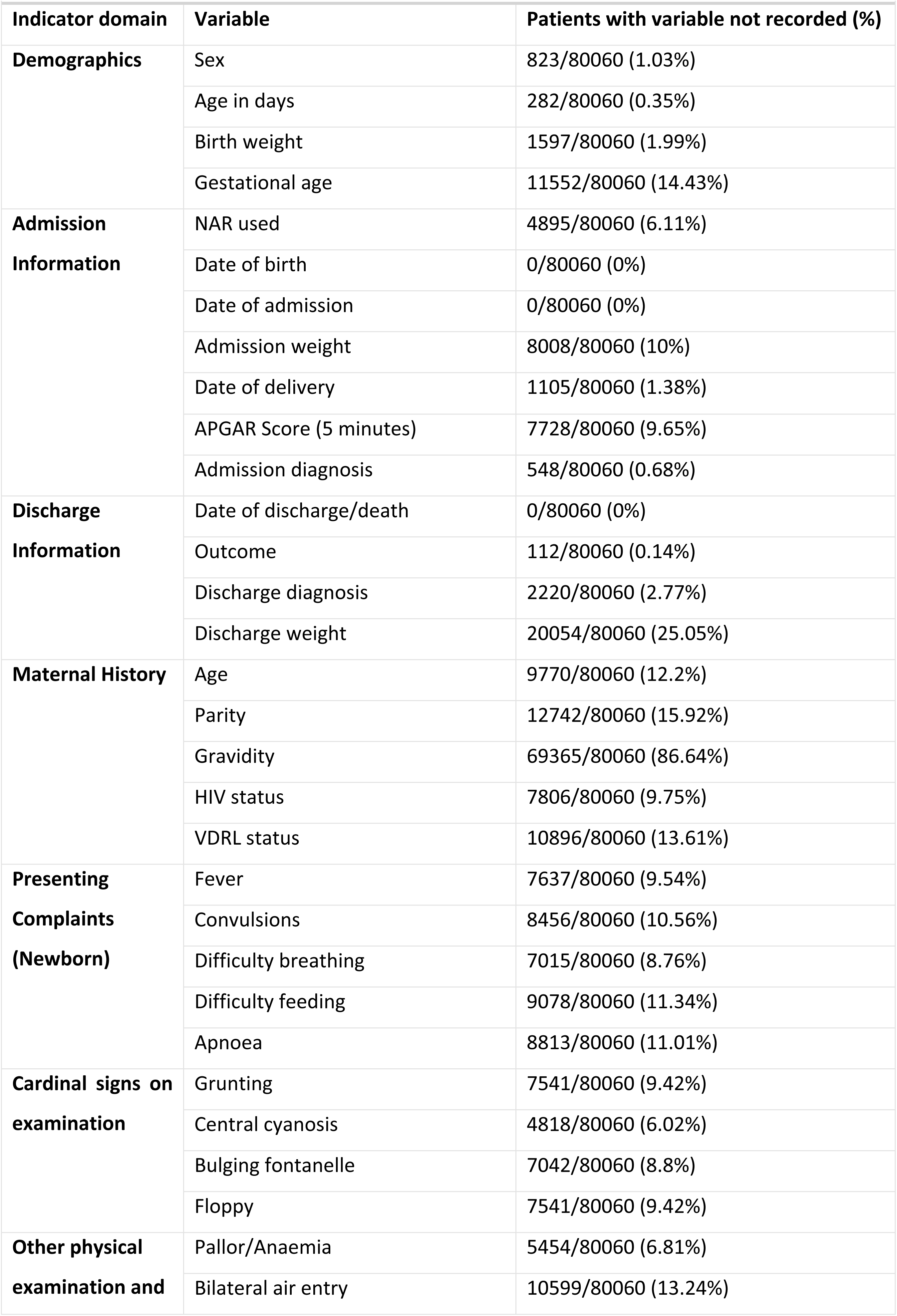
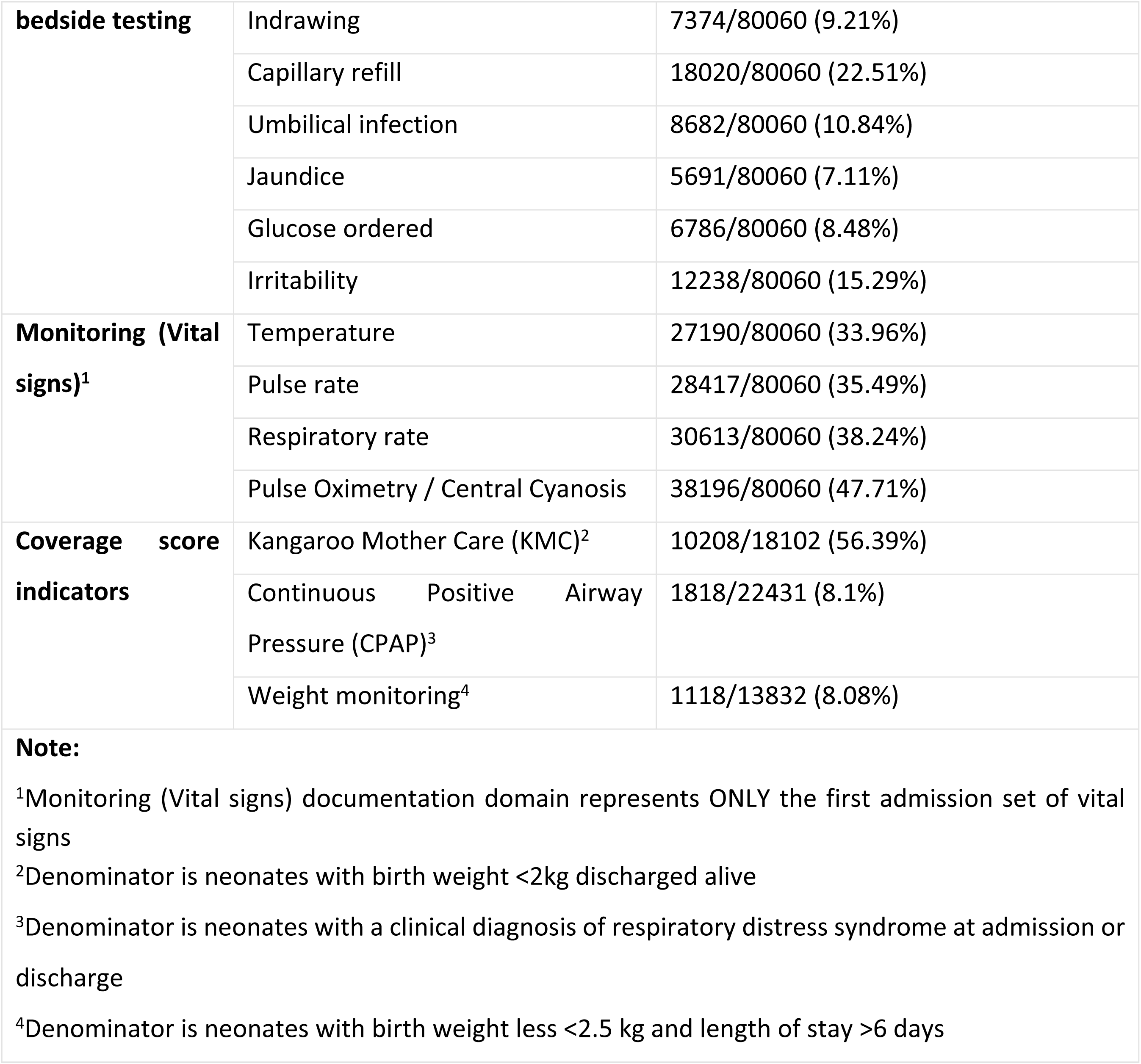
Proportion of records in which key items are not recorded

### Objective 1 Findings: Quality of documentation of in-patient neonatal care provided over time

Examining trends with data from across hospitals it can be seen that at the time all (new) hospitals joined the CIN-N, documentation completeness for 5/8 documentation domains was already around 80% or better (Figures 2, Table 4). This is likely attributable to most of these sites already using the NAR linked to being already part of CIN-Paediatrics [9, 14, 16]. Specifically, for *admission information*, *discharge information* and *demographics* documentation domains, performance was consistently >95%, with a median of >80% of patients having full documentation at admission (Table 4, Figure 2). For this reason, we do not further examine hospital specific trends for these domains.

**Figure 2:**
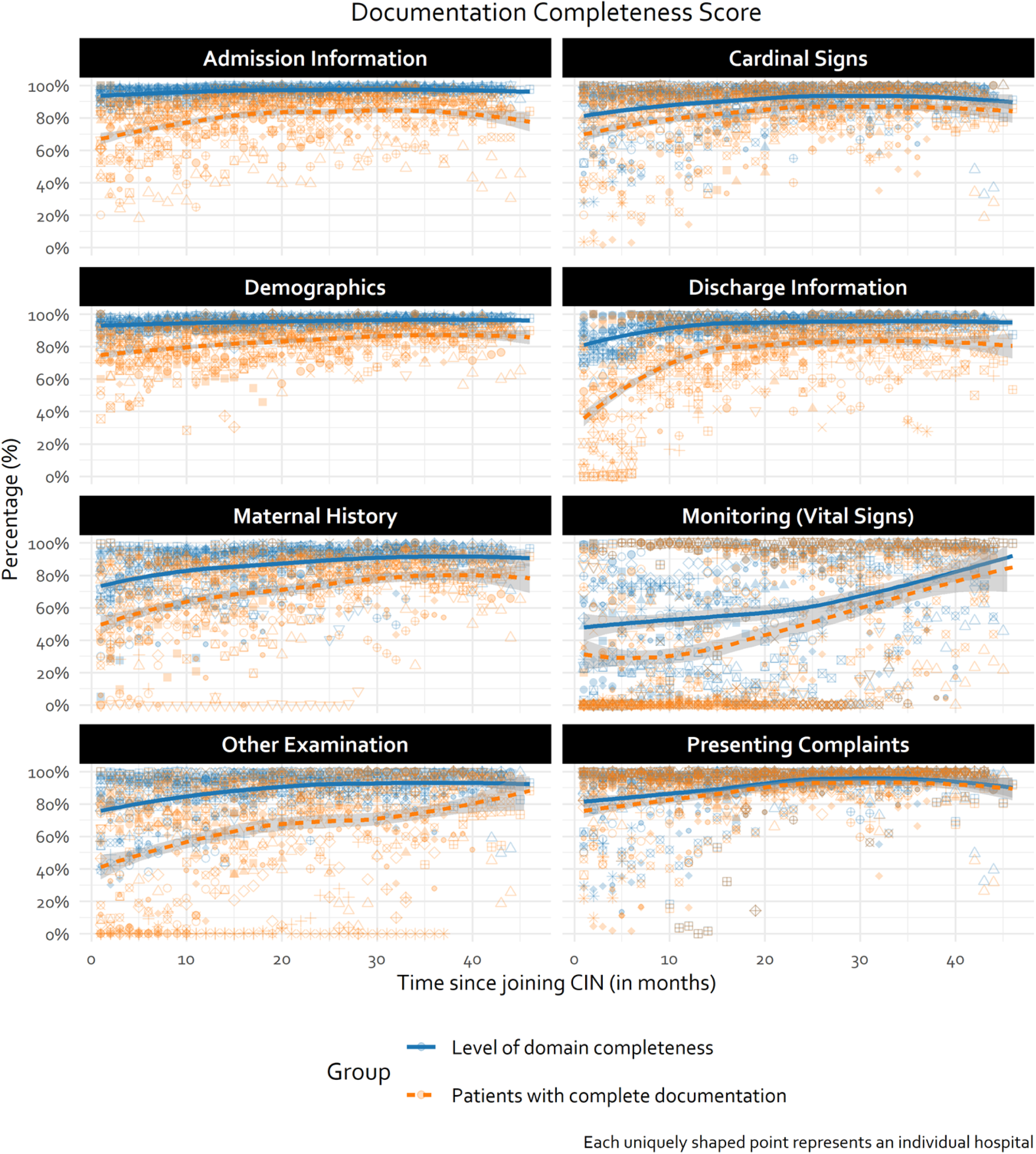
Domain-specific documentation trends over time. Domain completeness score summarised as an average of all individual patient domain-specific scores in each month. Trend line generated using LOWESS technique.

**Table 4:**
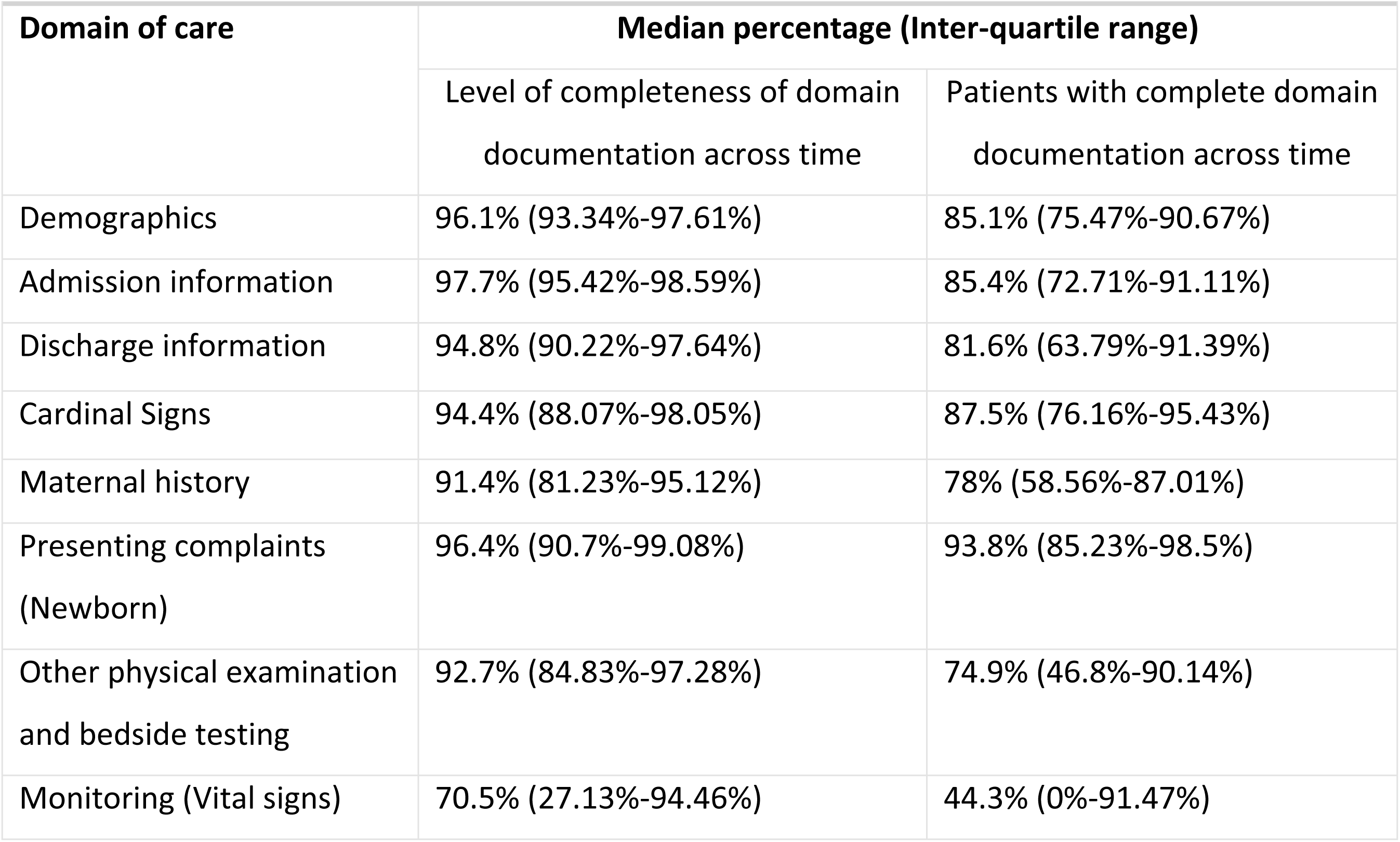
Domain documentation summary statistics pooled across all time periods (n= 80060 patients)

|  |  |  |
| --- | --- | --- |
| Domain of care | Median percentage (Inter-quartile range) |  |
|  | Level of completeness of domain documentation across time | Patients with complete domain documentation across time |
| Demographics | 96.1% (93.34%-97.61%) | 85.1% (75.47%-90.67%) |
| Admission information | 97.7% (95.42%-98.59%) | 85.4% (72.71%-91.11%) |
| Discharge information | 94.8% (90.22%-97.64%) | 81.6% (63.79%-91.39%) |
| Cardinal Signs | 94.4% (88.07%-98.05%) | 87.5% (76.16%-95.43%) |
| Maternal history | 91.4% (81.23%-95.12%) | 78% (58.56%-87.01%) |
| Presenting complaints<br>(Newborn) | 96.4% (90.7%-99.08%) | 93.8% (85.23%-98.5%) |
| Other physical examination<br>and bedside testing | 92.7% (84.83%-97.28%) | 74.9% (46.8%-90.14%) |
| Monitoring (Vital signs) | 70.5% (27.13%-94.46%) | 44.3% (0%-91.47%) |

For other domains, performance started lower with a suggestion from all hospitals’ data of improvement over time, but also considerable between-hospital variability e.g., *maternal history*, *other bedside examination,* and *monitoring* (*vital signs*) (Figure 2). We explore and demonstrate this considerable between hospital variability (e.g., *Other examination* domain) in Figure 3 (and Supplementary Figure 1). Plots display some examples of broad improvement (H13 and H20), some with static performance over time (e.g., H12) and some with rather erratic performance including occasional substantial declines (e.g., H2). Domains that often have lower baselines were those where documentation practices were less standardised prior to joining CIN-N. New information tools such as Transfer Forms (for sick newborns transferred from labour wards or theatres to NBU) or feedback on documentation of vital signs on NBU admission may have improved performance for *Maternal history* and *Monitoring (Vital signs)* domains across hospitals over time (Figures 2 and 3). However, the challenges with adoption or improvement are illustrated by both patterns where facilities starting very low showed gradual improvement and those starting high which either stagnated or got worse.

**Figure 3:**
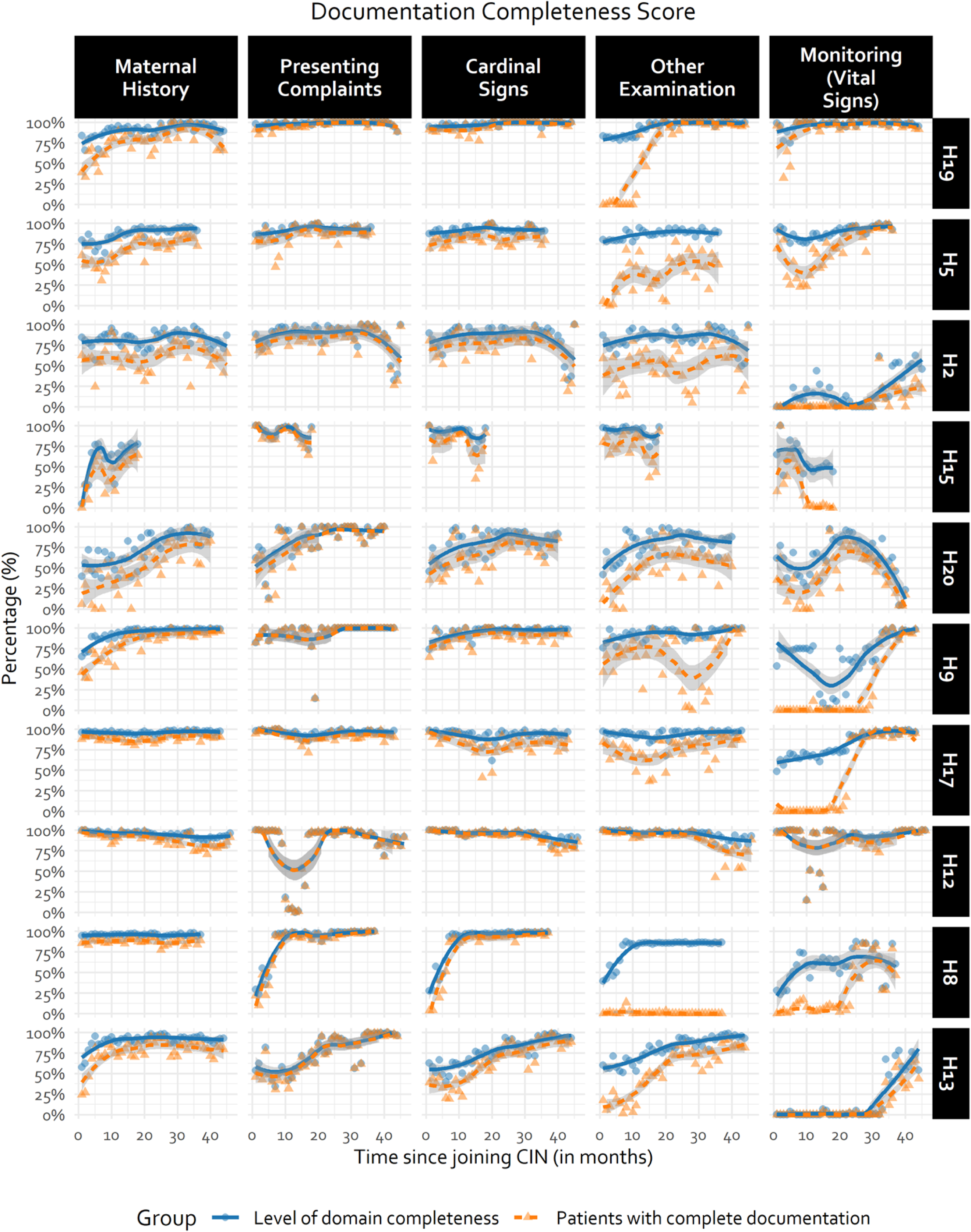
Illustration of hospital-specific documentation trends using a random selection of half the CIN-N hospitals. Hospital-specific trends for the remaining subset of hospitals can be found in Supplementary Figure 1. Trend line generated using LOWESS technique. Fewer observations in some hospitals due to different CIN-N joining dates.

All documentation domains demonstrated month-to-month improvements in the number of patients with complete domain documentation– even if modest in size – which are statistically significant (Table 5); In descending order, *monitoring (vital signs)*, *other physical examination and bedside testing* (i.e. Other Signs), *discharge information*, and *maternal history* domains demonstrated a statistically significant month-to-month relative improvement in number of patients with complete documentation of 7.6%, 2.9%, 2.4%, and 2.0% respectively (Table 5).

**Table 5:** Incidence rates of complete domain documentation at admission over time

|  |  |  |  |  |  |
| --- | --- | --- | --- | --- | --- |
| Count models with random intercept for hospitals and autocorrelation term for time respectively<sup>1</sup> |  |  |  |  |  |
| Domain | Term<sup>2</sup> | IRR | 95% CI | P-Value | ICC (Correlation)<sup>3</sup> |
| Admission Information | (Intercept) | 0.695 | 0.637 - 0.758 | <0.001 | 0.6682 (0.988) |
|  | Time | 1.006 | 1.004 - 1.007 | <0.001 |  |
| Cardinal signs on examination | (Intercept) | 0.665 | 0.573 - 0.772 | <0.001 | 0.66 (0.943) |
|  | Time | 1.007 | 1.002 - 1.012 | 0.009 |  |
| Demographics | (Intercept) | 0.755 | 0.716 - 0.796 | <0.001 | 0.6848 (0.976) |
|  | Time | 1.004 | 1.002 - 1.005 | <0.001 |  |
| Discharge Information<sup>4</sup> | (Intercept) | 0.344 | 0.258 - 0.458 | <0.001 | 0.0239 (0.954) |
|  | Time | 1.024 | 1.014 - 1.034 | <0.001 |  |
| Maternal History | (Intercept) | 0.373 | 0.254 - 0.547 | 0.004 | 0.5341 (0.984) |
|  | Time | 1.020 | 1.01 - 1.03 | 0.022 |  |
| Monitoring (Vital signs)<sup>4</sup> | (Intercept) | 0.028 | 0.01 - 0.078 | <0.001 | 0.5121 (0.982) |
|  | Time | 1.076 | 1.046 - 1.107 | 0.006 |  |
| Other physical examination and bedside testing<sup>4</sup> | (Intercept) | 0.206 | 0.11 - 0.388 | <0.001 | 0.5064 (0.991) |
|  | Time | 1.029 | 1.016 - 1.043 | <0.001 |  |
| Presenting Complaints | (Intercept) | 0.711 | 0.618 - 0.819 | <0.001 | 0.6752 (0.905) |
|  | Time | 1.008 | 1.002 - 1.013 | 0.006 |  |
| <sup>1</sup> Time treated as continuous covariate representing consecutive months in CIN |  |  |  |  |  |
| <sup>2</sup>Intercept term provides baseline rate at month 1 of joining the CIN. |  |  |  |  |  |
| <sup>3</sup>Intra-Cluster Correlation (ICC), with autocorrelation parameter provided in the brackets. The choice to account for heterogeneity between hospitals while allowing for varying effects of time (i.e., a random slope) within each hospital was more suitable that just a hospitals random intercepts model. However, a hospitals random intercept model with an autocorrelation term for time outperformed a random slopes model equivalent (Supplementary Table 2). |  |  |  |  |  |
| <sup>4</sup>Used negative binomial models in place of Poisson models given the equidispersion assumption violation (Supplementary Table 2) |  |  |  |  |  |

At the time of joining CIN-N, less than 50% of the patients admitted to the hospitals had complete documentation of *Discharge information*, *Monitoring (Vital signs)*, *Other physical examination and bedside testing*, and *Maternal history* domains, with a substantive amount of variance in the outcome explained by hospital factors as illustrated by the high ICCs (Table 5, Figure 3). Hospitals with higher baseline performance tended to demonstrate slower rates of improvement than hospitals with lower baseline performance as illustrated in Figure 3 (Table 5, H13 versus H17 in Figure 3).

### Objective 2 Findings: Accuracy of essential neonatal intervention prescriptions over time

Given the good quality of prescribing data from CIN-N and the reasonable assumptions about the meaning of missing prescribing data, treatment prescribing accuracy was evaluated for the common antibiotics, feeds, and fluids in NBUs. Domain specific treatment accuracy scores revealed an increasing proportion of patients with accurate fluids and feeds prescriptions from approximately 40% to 60%, and 15% to 40% respectively, although feeds prescribing accuracy then regresses to 25% (Figure 4).

**Figure 4:**
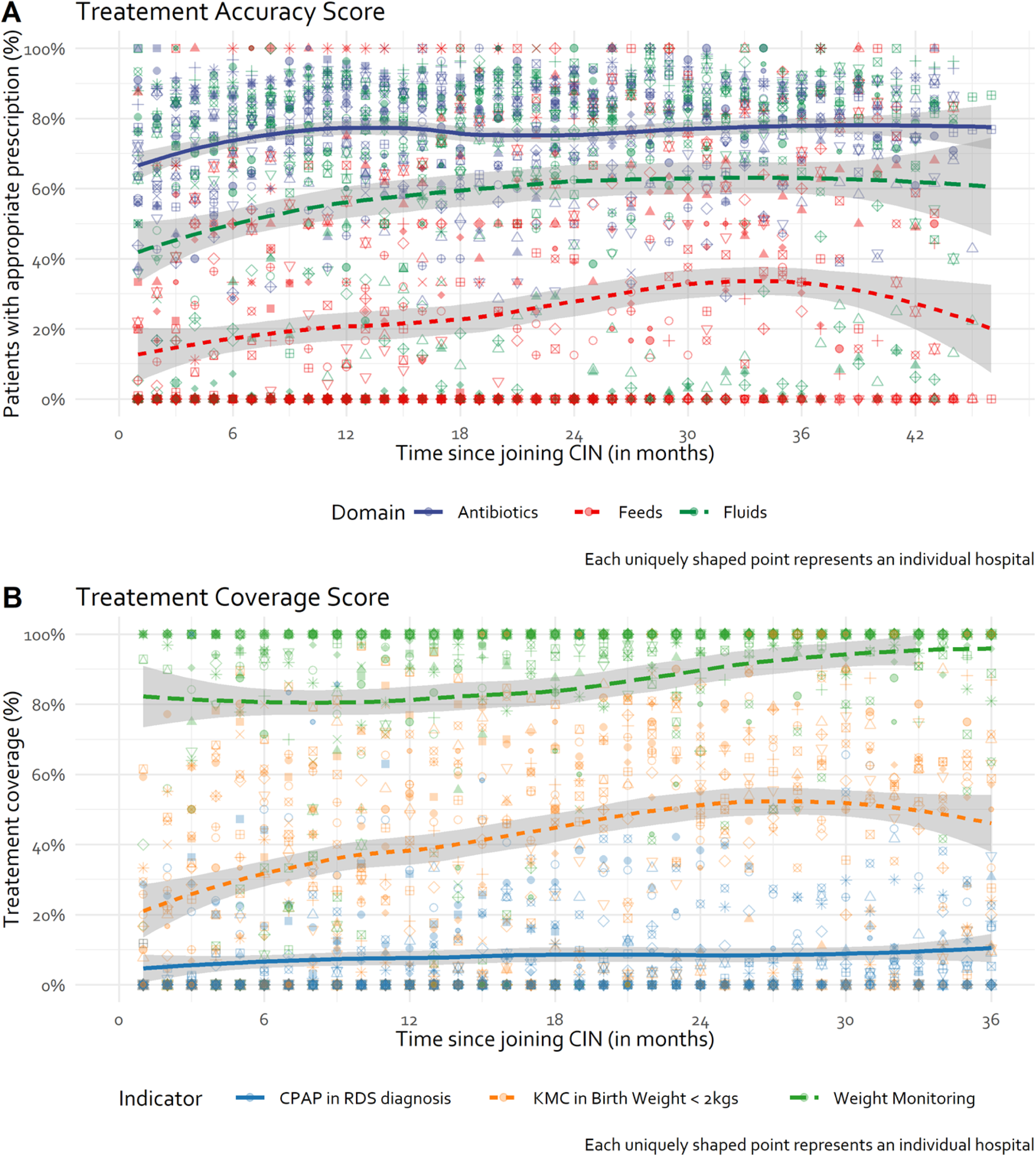
Overall trend in treatment accuracy and coverage. KMC: Kangaroo Mother Care; CPAP: Continuous Positive Airway Pressure; RDS: Respiratory Distress Syndrome. Trend line generated using LOWESS technique.

Antibiotic prescription shows a modest improvement in accuracy from around 65% to 80% within the first 12 months, after which it then fluctuated around 80% over time across all CIN-N admissions. Treatment coverage levels for KMC demonstrated an increase over time from 20% to 40% in neonates with birth weight <2kg (Figure 4). There was a small increase in CPAP coverage levels over time in neonates with a clinical diagnosis of respiratory distress syndrome (RDS) from 4% to 10%. Repeated weight monitoring for sick neonates improved from 80% to approximately 95% over time (Figure 4). There is evidence of moderate to high hospital variability in both treatment accuracy and coverage scores in CIN-N hospitals (Figure 4).

While antibiotic treatment accuracy seems to have a ceiling effect of 80% in pooled hospital data, hospital specific plots (Figure 5, Supplementary Figure 2) indicate that hospitals can consistently attain accuracy levels > 80% (H8, H11, H19), suggesting improvements in other sites would be possible. Hospital-specific trends suggest fluids prescribing accuracy improves from a lower baseline for some hospitals (e.g., H5, H12, H20) but there is still a long way to go, with considerable between hospital variability evident over time (Figure 5, Supplementary Figure 2). Similarly, feeds prescribing accuracy shows some improvement in some hospitals from a lower baseline (e.g., H5, H8) but in others performance is erratic (H2, H12), with most performing consistently poorly over time (e.g., H13, H19) (Figure 5, Supplementary Figure 2).

**Figure 5:**
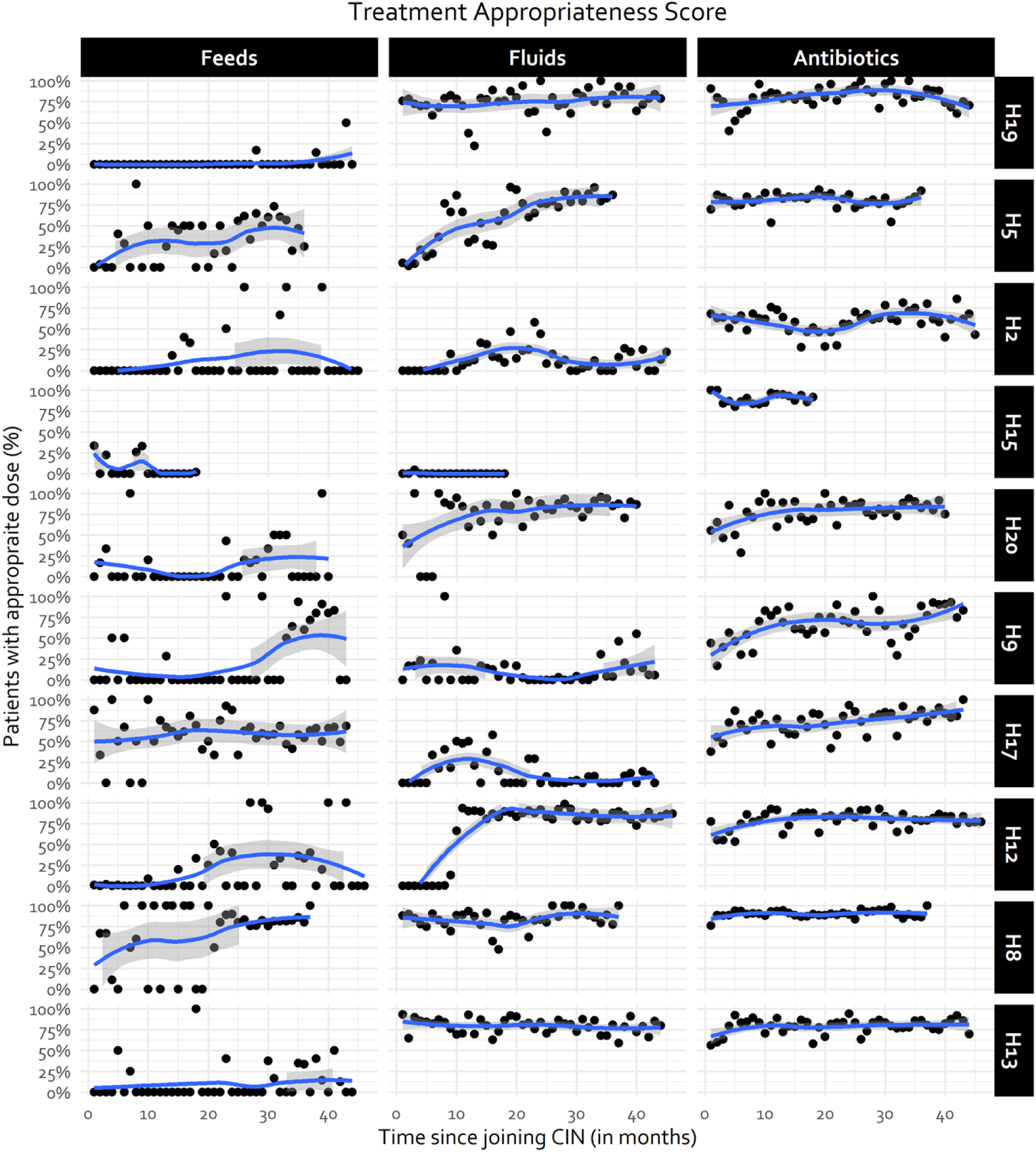
Hospital-specific treatment accuracy trends for half of randomly selected CIN-N hospitals. Hospital-specific trends for the remaining subset of hospitals can be found in Supplementary Figure 2. Trend line generated using LOWESS technique. Fewer observations in some hospitals due to different CIN-N joining dates.

On average, 73.5%, 10.8% and 22.8% of the patients in the CIN-N received the correct antibiotic, feeds, and fluids treatment at the time when hospitals joined the CIN-N (Table 6). There was a modest statistically significant month-to-month relative increase in correct inpatient treatment after joining the CIN of 2.8% and 1.4% for feeds and fluids prescribing accuracy. Antibiotic prescriptions showed no statistically significant month-to-month improvement after joining CIN-N (Table 6). The high ICC from the antibiotics and fluids mixed models suggests that much of the variation in the prescribing practices accuracy is associated with hospital-level factors.

**Table 6:**
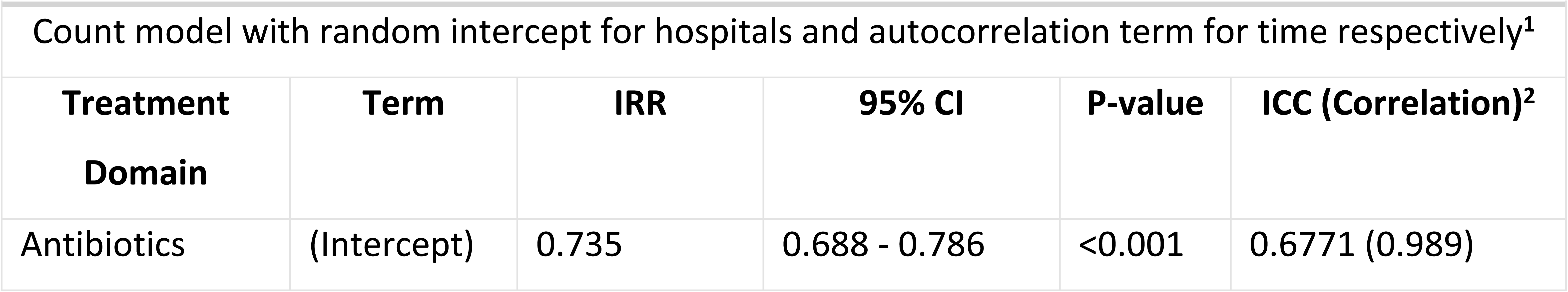
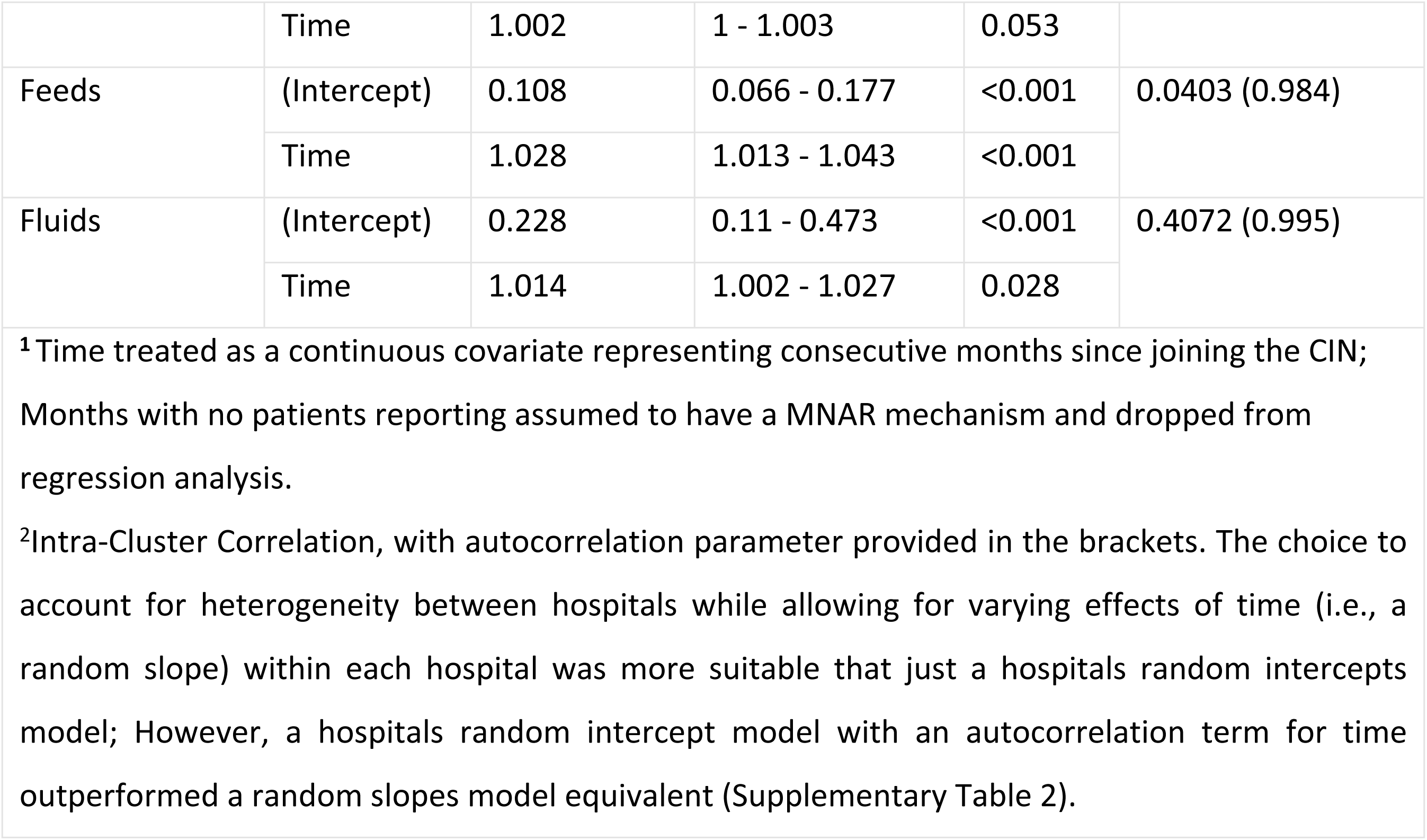
Incidence rates of correct treatment prescribing at admission over time

### Sensitivity analyses findings

It was reasonable to use Poisson models (which assumes equidispersion) in all but four documentation domains (*Discharge information, Monitoring (Vital Signs), Other physical examinations and bedside testing (i.e., Other Signs)* documentation domains) and the *Feeds* treatment accuracy domain which showed evidence suggestive of overdispersion (Supplementary Table 2). Where the equidispersion assumption was violated, negative binomial models were used and informed any inference drawn.

## Discussion

### Summary of findings

This study aimed at determining if quality routine clinical data might be generated from CIN-N hospitals and if the quality of data improved over time. As the data quality was reasonable, they were then used to determine whether essential treatments or interventions are being correctly prescribed to newborns and to track intervention adoption. From the time hospitals joined the CIN-N, around 80% of newborns had complete documentation in 5/8 documentation domains (Table 4). This relatively good performance at baseline may be a consequence of participation in the paediatric CIN by most of these hospitals prior to formal extension of CIN to NBUs (i.e., CIN-N) with many paediatric practitioners previously exposed to use of the NAR, ETAT+ training and national neonatal guidelines [13]. All documentation domains demonstrated month-to-month statistically significant albeit modest improvements (between 0.6% and 7.6% per month) in the number of patients with complete domain documentation (Figure 4, Table 5). On average, 73.5%, 10.8% and 22.8% of the newborns with treatment orders in the CIN-N for first-line antibiotics, feeds, and fluids had correct prescriptions at the time when hospitals joined the CIN-N (Table 6). There is a modest statistically significant 2.8% and 1.4% month-to-month relative increase in accurate feeds and fluids prescription after joining the CIN-N resulting in an end line performance of around 40% and 60% respectively. Antibiotic prescribing showed no statistically significant month-to-month change.

Although sometimes modest the improvements observed were often sustained during the COVID-19 pandemic (perhaps with the exception of feed prescribing) that restricted network engagement activities to brief online meetings between April 2020 and December 2021. Improvements also occurred over a period of 4 years during which junior medical staff on NBUs changed every 3 months with frequent changes also in senior staff [19]. Across the entire period CIN-N sustained distribution of feedback reports and the magnitude of improvements observed are in keeping with findings from many audit and feedback interventions [32].

Coverage levels for KMC in neonates with birth weight <2kg and CPAP in neonates with clinically diagnosed respiratory distress syndrome (RDS) demonstrated an increase over time from 20% to 40% and 4%-10%; repeated weight monitoring for sick neonates with birth weight <2.5 kg and length of stay >6 days improved from 80% to approximately 95% over time (Figure 4). There was evidence of moderate to high hospital variability in documentation, treatment accuracy, and coverage scores in CIN-N hospitals; as shown previously hospitals with higher baseline performance evidence slower rates of improvement than hospitals with lower baseline performance in some cases perhaps linked to ceiling effects [33].

### Comparison to other findings

Previous studies in Kenya depict a health system that is struggling to collect quality data that is usable for decision making especially for neonatal care [18, 34]. The poor quality of neonatal clinical data has been widely reported in other African countries [35, 36], this undermines efforts to track the scale up of quality care [37, 38]. Implementation of a learning health system across hospitals utilising a common data platform to facilitate routine audit and feedback cycles have been shown to improve the documentation of patient data and its subsequent use in care improvement [9, 37, 39]. Employing findings, tools and practices from previous studies and progressively engaging more hospitals we demonstrate that data can be collected using a common data platform as part of a learning health system approach from a network of hospitals’ NBUs; we further show that these data can be useful for identifying potential gaps in care (e.g., treatment accuracy) with an aim of improving the quality of care provided in facilities and tracking outcomes at scale [13-16, 18, 40]. To our knowledge this is the largest reported long-term neonatal learning health system platform in SSA, serving as an exemplar actionable health information system in line with WHO standards [13, 15, 16, 41].

Findings from scoping reviews suggest that having better data can help improve quality of care if coupled with development of local leadership, training, and use of local improvement strategies such as mortality audits or quality improvement cycles; This can help reduce inpatient neonatal mortality in low-income country hospitals [42–45]. However, the complex intervention strategies required to tackle multiple quality and safety concerns in hospitals may make it challenging to demonstrate mortality reductions over the short term [46]. High-quality data platforms may therefore be especially helpful to track whether hospital quality of care and mortality rates are improving over the long-term. Hospital neonatal outcomes may also be influenced by the successful scaling up of key interventions such as CPAP and KMC. It is therefore essential to be able to track their adoption at scale. However, outside specific research studies these data are rarely reported and the effects of programmes supporting such scaling up therefore remain largely unknown. The CIN-N data platform we describe, by spanning aspects of care rarely included in other LMIC quality assessment approaches, offers one means to track adoption over the long-term and provides a hypothesis generating platform for implementation research linked to observed variations in quality of care and intervention rollout [42, 47, 48].

### Implications of findings

For key clinical data domains (i.e., demographic, admission, and discharge information) there was good data documentation at the time hospitals joined the CIN-N. This is likely attributable to most of these sites being already part of CIN-Paediatrics, where organisationally, the learning health system culture was already being cultivated, allowing these hospitals to take advantage of the roll-out and dissemination of tools like the NAR coupled with ETAT+ training. This also does suggest that it is possible to introduce tools that capture essential clinical data with missingness rates of 20% or less in routine SSA hospital settings. Analyses of such data do then need to account for missingness using appropriate statistical techniques to reduce potential biases [49, 50]. The slow but steady month-to-month improvement illustrates how long it takes to change clinical behaviours for some forms of patient documentation.

For several documentation domains (e.g., *Other physical examination, Maternal History, Monitoring Vital Signs*) and all treatment accuracy and coverage domains except antibiotic prescribing accuracy, considerable variability in performance between hospitals remains a persistent challenge. In some cases, this may reflect a “ceiling” effect (e.g., *Fluids* prescribing accuracy). It is evident, however, that some hospitals can attain higher accuracy levels consistently (Figures 3 and 5), suggesting improvements in other sites would be possible. Similarly, trends in accuracy in the prescription of feeds and fluids are quiet erratic and vary within and between hospitals (Figure 6). These erratic patterns might have been exacerbated by the limited interaction of hospitals through CIN meetings during the COVID-19 pandemic and other challenges to sustaining quality care such as human resource shortages and labour strikes [19, 20]. Learning from ‘positive deviants’ may be informative. Prior research suggests good performers have adequate and supportive staffing, participate actively in local clinical audits and feedback process, and have good supervision by unit leads [14–16, 51].

Better theory-driven ways of conducting audit and feedback might be required within CIN-N to improve quality of care and treatment accuracy. [32] For example, more active feedback might be needed for more complex tasks such as to promote accurate prescribing. Further elaborations might explicitly address (1) capacity limitations of CIN-N hospitals and clinical teams to produce the improvements required, (2) lessons learned about the identity and culture of each individual CIN-N hospital and site specific barriers to change (3) specific use of behavioural thinking that directly supports positive clinical behaviours by ensuring feedback is actionable, controllable and timely [39, 52].

### Strengths and limitations

This study is among the few in SSA focusing on documentation of new-born data, an implementation of strategic objective 2 and 5 of the Every Newborn Action Plan [6, 53, 54]. The CIN-N generates data from more than 20 NBUs across Kenya by improving routine data sources in a strategy that is relatively low-cost and scalable as the central data management and data quality assurance processes involved can benefit from economies of scale [9, 55]. However, the data generated are limited only to ‘documentation’, limiting the range of measures of quality e.g., whether prescriptions were correct. Key limitations therefore remain such as confirmation of whether treatment was dispensed as prescribed (i.e., treatment adherence), and, for interventions such as CPAP, it can be hard to determine the best denominator population which would ideally be newborns that might have benefited from its use. This problem in identifying suitable denominators that enable evaluation of the appropriate use of interventions mean tracking adoption may frequently be based on a cruder measure of frequency of (documented) use. Furthermore, for some indicators applicable to only relatively small numbers of patients performance may appear erratic because there are few data points per month. Thus, less frequent monitoring over longer time periods may be required to sensibly track trends.

## Conclusions

It is possible to introduce tools that capture essential clinical data often in 80% or more of newborns admitted to routine SSA hospital settings engaged in a centrally supported peer-to-peer network, but analyses of such data need to account for missingness using appropriate statistical techniques. Improvements in quality indicators are on average modest but valuable on a month-to-month basis and occur over a prolonged period that included the COVID pandemic. Average effects mask considerable temporal and between hospital variability with some hospitals demonstrating high levels of performance for indicators likely to be important to patient safety and outcomes such as feeding or antibiotic treatment prescribing accuracy. Learning from high-performing hospitals and continuously deploying better theory-driven feedback approaches may help realise better improvements more widely. However, considerable system challenges such as rapid staff turnover, general staff shortages and ongoing material resource challenges likely contribute to persistent problems delivering quality care. Such quality clinical data (and associated platforms) can support better impact evaluation, performance benchmarking, exploration of links between health system inputs and outcomes and critical scrutiny of geographic variation in quality and outcomes of hospital care [56]. Efforts to improve the quality of clinical data from SSA are needed to support these objectives remain much needed.

## Declarations

## Abbreviations

A&F: Audit and Feedback
CIN: Clinical Information Network
CPAP: Continuous Positive Airway Pressure
ETAT+: Emergency Treatment and Triage plus admission
KEMRI: Kenya Medical Research Institute
KMC: Kangaroo Mother Care
KPA: Kenya Paediatric Association
LMICs: Low and middle-income countries
MoH: Ministry of Health
NAR: Neonatal Admission Record
NBU: Newborn Unit
SERU: KEMRI’s Scientific and Ethics Review Unit
SSA: Sub-Saharan Africa
WHO: World Health Organization.

## Ethics approval and consent to participate

Ethical approval was provided by the KEMRI Scientific and Ethical Review Committee (SERU 3459). Individual patient consent for the de-identified clinical data was judged to not be required, but consent from participating hospitals was sought.

## Consent for publication

This protocol is published with the permission of the Director of Kenya Medical Research Institute (KEMRI). At this stage with no protocol containing no data from any individual person, individual consent is not applicable.

## Availability of Data and Materials

The datasets generated and/or analysed during the current study are not publicly available due to the primary data being owned by the hospitals and their counties with the Ministry of Health; The research staff do have permission to share the data without further written approval from both the KEMRI-Wellcome Trust Data Governance Committee and the Facility, County or Ministry of Health as appropriate to the data request.

Requests for access to primary data from qualitative research by people other than the investigators will be submitted to the KEMRI-Wellcome Trust Research Programme data governance committee as a first step through, who will advise on the need for additional ethical review by the KEMRI Research Ethics Committee.

## Author contributions

**Conceptualization:** TT, ME

**Data Curation:** GM, LM, KW, JW, DM

**Formal Analysis:** TT, JA, DC, MM, ME

**Methodology:** TT, JA, ME

**Investigation:** TT, JW, DC, JA, ME

**Software:** TT, JW, KW, LM, GM

**Supervision:** ME

**Writing – Original Draft Preparation:** TT

**Writing – Review & Editing:** TT, DC, JA, JW, GM, KW, LM, DM, GI, MM, ME

All authors revised the manuscript, approved the final version and agreed to be accountable for the findings.

## Data Availability

The datasets generated and/or analysed during the current study are not publicly available due to the primary data being owned by the hospitals and their counties with the Ministry of Health The research staff do have permission to share the data without further written approval from both the KEMRI-Wellcome Trust Data Governance Committee and the Facility, County or Ministry of Health as appropriate to the data request. Requests for access to primary data from qualitative research by people other than the investigators will be submitted to the KEMRI-Wellcome Trust Research Programme data governance committee as a first step through, who will advise on the need for additional ethical review by the KEMRI Research Ethics Committee.

## Acknowledgements

The **Clinical Information Network (CIN) Group**: The CIN group hospital teams who are tagged to collaborate in the network’s development, data collection, data management, implementation of audit and feedback interventions and who will participate in this study include the following focal persons:

a. **Paediatricians:** Juma Vitalis, Nyumbile Bonface, Roselyne Malangachi, Christine Manyasi, Catherine Mutinda, David Kibiwott Kimutai, Rukia Aden, Caren Emadau, Elizabeth Atieno Jowi, Cecilia Muithya, Charles Nzioki, Supa Tunje, Dr. Penina Musyoka, Wagura Mwangi, Agnes Mithamo, Magdalene Kuria, Esther Njiru, Mwangi Ngina, Penina Mwangi, Rachel Inginia, Melab Musabi, Emma Namulala, Grace Ochieng, Lydia Thuranira, Felicitas Makokha, Josephine Ojigo, Beth Maina, Catherine Mutinda, Mary Waiyego, Bernadette Lusweti, Angeline Ithondeka, Julie Barasa, Meshack Liru, Elizabeth Kibaru, Alice Nkirote Nyaribari, Joyce Akuka, Joyce Wangari;
b. **Nurses:** Amilia Ngoda, Aggrey Nzavaye Emenwa, Patricia Nafula Wesakania, George Lipesa, Jane Mbungu, Marystella Mutenyo, Joyce Mbogho, Joan Baswetty, Ann Jambi, Josephine Aritho, Beatrice Njambi, Felisters Mucheke, Zainab Kioni, Jeniffer, Lucy Kinyua, Margaret Kethi, Alice Oguda, Salome Nashimiyu Situma, Nancy Gachaja, Loise N. Mwangi, Ruth Mwai, irginia Wangari Muruga, Nancy Mburu, Celestine Muteshi, Abigael Bwire, Salome Okisa Muyale, Naomi Situma, Faith Mueni, Hellen Mwaura, Rosemary Mututa, Caroline Lavu, Joyce Oketch, Jane Hore Olum, Orina Nyakina, Faith Njeru, Rebecca Chelimo, Margaret Wanjiku Mwaura, Ann Wambugu, Epharus Njeri Mburu, Linda Awino Tindi, Jane Akumu, Ruth Otieno, Slessor Osok;
c. **Health Record Information Officers (HRIOs):** Seline Kulubi, Susan Wanjala, Pauline Njeru, Rebbecca Mukami Mbogo, John Ollongo, Samuel Soita, Judith Mirenja, Mary Nguri, Margaret Waweru, Mary Akoth Oruko, Jeska Kuya, Caroline Muthuri, Esther Muthiani, Esther Mwangi, Joseph Nganga, Benjamin Tanui, Alfred Wanjau, Judith Onsongo, Peter Muigai, Arnest Namayi, Elizabeth Kosiom, Dorcas Cherop, Faith Marete, Johanness Simiyu, Collince Danga, Arthur Otieno Oyugi, Fredrick Keya Okoth.

The **Clinical Information Network (CIN) Group**’s monitored email address is and the list can change when new paediatrician(s), nurse(s) or HRIO leave or come into the hospital.

## Open access

This is an open access article distributed in accordance with the Creative Commons Attribution 4.0 Unported (CC BY 4.0) license, which permits others to copy, redistribute, remix, transform and build upon this work for any purpose, provided the original work is properly cited, a link to the licence is given, and indication of whether changes were made.

## Appendix

**Supplementary Table 1:**
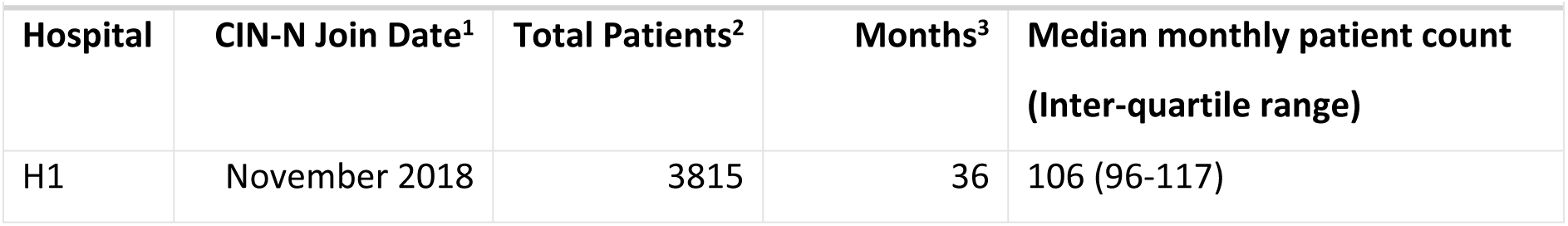
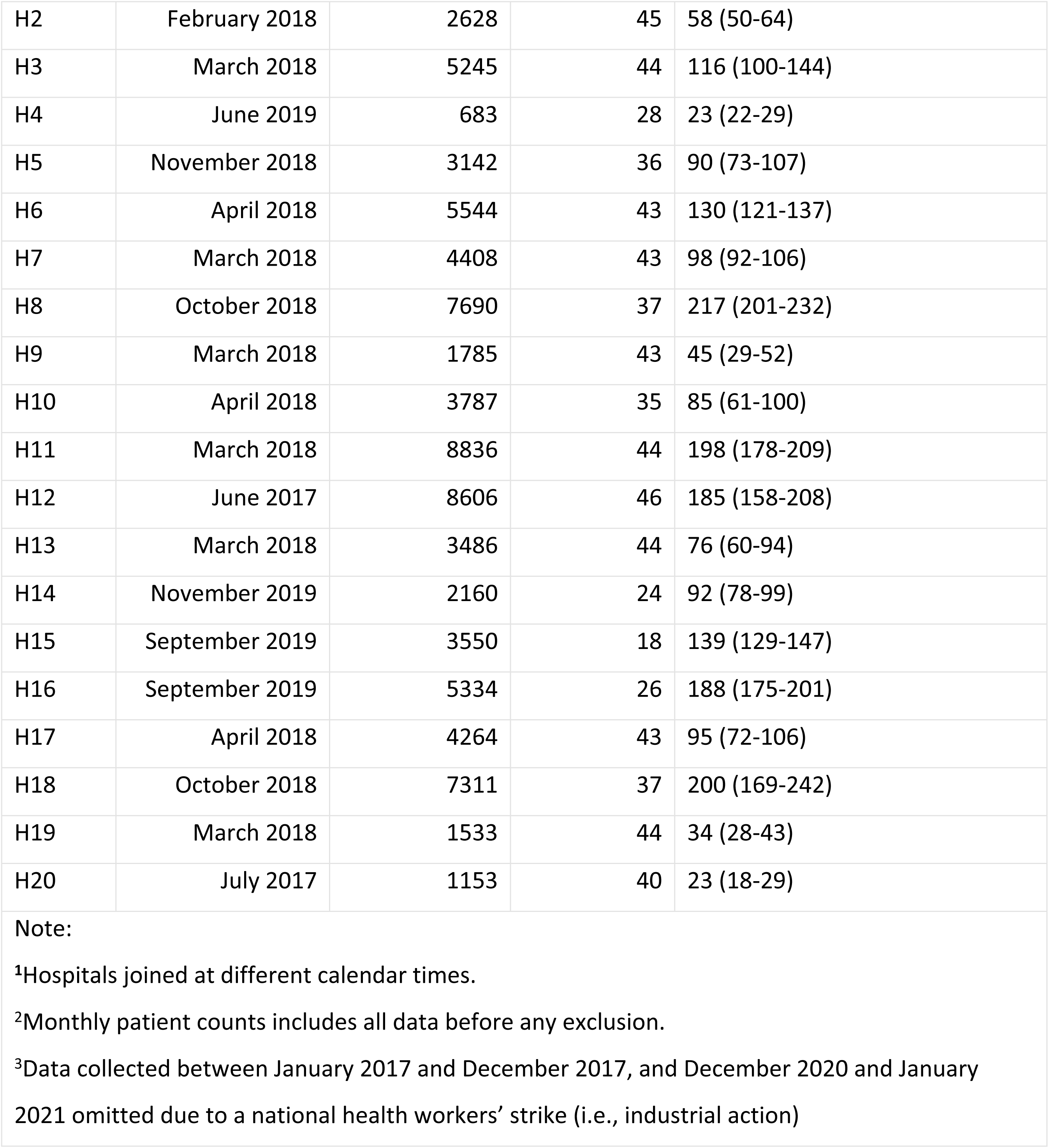
Hospitals’ CIN-N membership and patient volumes

| Supplementary Table 1: Hospitals' CIN-N membership and patient volumes |  |  |  |  |
| --- | --- | --- | --- | --- |
| Hospital | CIN-N Join Date<sup>1</sup> | Total Patients<sup>2</sup> | Months<sup>3</sup> | Median monthly patient count<br>(Inter-quartile range) |
| H1 | November 2018 | 3815 | 36 | 106 (96-117) |
| H2 | February 2018 | 2628 | 45 | 58 (50-64) |
| H3 | March 2018 | 5245 | 44 | 116 (100-144) |
| H4 | June 2019 | 683 | 28 | 23 (22-29) |
| H5 | November 2018 | 3142 | 36 | 90 (73-107) |
| H6 | April 2018 | 5544 | 43 | 130 (121-137) |
| H7 | March 2018 | 4408 | 43 | 98 (92-106) |
| H8 | October 2018 | 7690 | 37 | 217 (201-232) |
| H9 | March 2018 | 1785 | 43 | 45 (29-52) |
| H10 | April 2018 | 3787 | 35 | 85 (61-100) |
| H11 | March 2018 | 8836 | 44 | 198 (178-209) |
| H12 | June 2017 | 8606 | 46 | 185 (158-208) |
| H13 | March 2018 | 3486 | 44 | 76 (60-94) |
| H14 | November 2019 | 2160 | 24 | 92 (78-99) |
| H15 | September 2019 | 3550 | 18 | 139 (129-147) |
| H16 | September 2019 | 5334 | 26 | 188 (175-201) |
| H17 | April 2018 | 4264 | 43 | 95 (72-106) |
| H18 | October 2018 | 7311 | 37 | 200 (169-242) |
| H19 | March 2018 | 1533 | 44 | 34 (28-43) |
| H20 | July 2017 | 1153 | 40 | 23 (18-29) |

Note:
<sup>1</sup>Hospitals joined at different calendar times.
<sup>2</sup>Monthly patient counts includes all data before any exclusion.
<sup>3</sup>Data collected between January 2017 and December 2017, and December 2020 and January 2021 omitted due to a national health workers' strike (i.e., industrial action)

**Supplementary Table 2:** Sensitivity analysis evaluating if there is any evidence that negative binomial model assumptions might have been violated?

|  |  |  |  |  |  |  |  |  |
| --- | --- | --- | --- | --- | --- | --- | --- | --- |
| Domain<sup>1</sup> | Over-dispersion value<sup>2</sup> | Equi-dispersion LRT p-value<sup>2</sup> | Nested model LRT p-value<sup>3</sup> | Auto-correlation LRT p-value<sup>4</sup> | AIC | BIC | Log likelihood | Deviance |
| Admission Information | 0 | 1 | <0.001 | <0.001 | 4988.35 | 5015.89 | -2488.18 | 4976.35 |
| Cardinal Signs | 0.05 | 0.581 | <0.001 | <0.001 | 6071.43 | 6099.19 | -3029.71 | 6059.43 |
| Demographics | 0 | 0.915 | <0.001 | <0.001 | 5187.99 | 5215.76 | -2588 | 5175.99 |
| Discharge Information | 0.3 | 0.03 | <0.001 | <0.001 | 6363.95 | 6391.72 | -3175.97 | 6351.95 |
| Maternal History | 0.117 | 0.303 | <0.001 | <0.001 | 6012.4 | 6040.17 | -3000.2 | 6000.4 |
| Monitoring (Vital Signs) | 2.254 | <0.001 | <0.001 | <0.001 | 5124.04 | 5151.66 | -2556.02 | 5112.04 |
| Other Examinations | 2.442 | <0.001 | <0.001 | <0.001 | 6176.34 | 6204.11 | -3082.17 | 6164.34 |
| Presenting Complaints | 0 | 0.902 | <0.001 | <0.001 | 6291.42 | 6319.18 | -3139.71 | 6279.42 |
| Antibiotics | 0 | 0.945 | 0.097 | <0.001 | 4477.24 | 4504.85 | -2232.62 | 4465.24 |
| Feeds<sup>4</sup> | 0.571 | 0.001 | <0.001 | <0.001 | 2263.05 | 2289.49 | -1125.52 | 2251.05 |
| Fluids | 0 | 1 | <0.001 | <0.001 | 4171.96 | 4199.22 | -2079.98 | 4159.96 |

**Note:**
<sup>1</sup>Evidence from all the final documentation completeness and treatment appropriateness negative binomial and Poisson models fitted (Tables 5 and 6) is suggestive that the model errors are normally distributed (i.e., the conditional outcomes were normally distributed) and log-linear (Supplementary Figures 3-5) thus suitable for these analyses.
<sup>2</sup>Model comparison test between negative binomial- and Poisson model structure for evidence in supporting presence of overdispersion.
<sup>3</sup>Comparison test between random slope-intercept model and random intercept only models for evidence in support of allowing time to vary within hospitals
<sup>4</sup>Comparison between random slope-intercept model (for time and hospital respectively) and random intercept model with auto-correlation term for time for exploring how best to allow time to vary within hospitals during model specification.

**Supplementary Figure 1:**
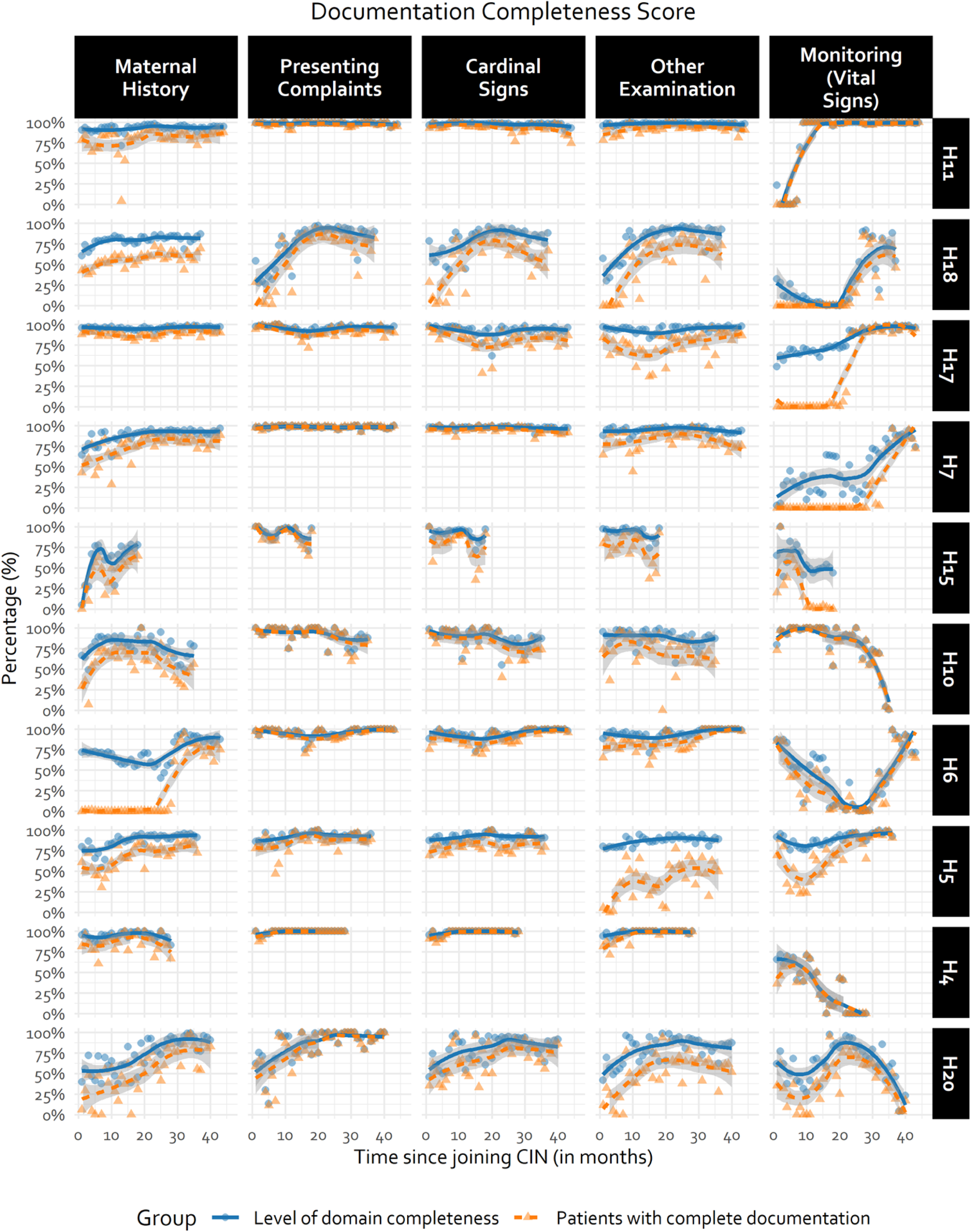
Hospital-specific documentation trends for last half of randomly selected CIN-N hospitals. Trend line generated using LOWESS technique. Fewer observations in some hospitals due to different CIN-N joining dates.

**Supplementary Figure 2:**
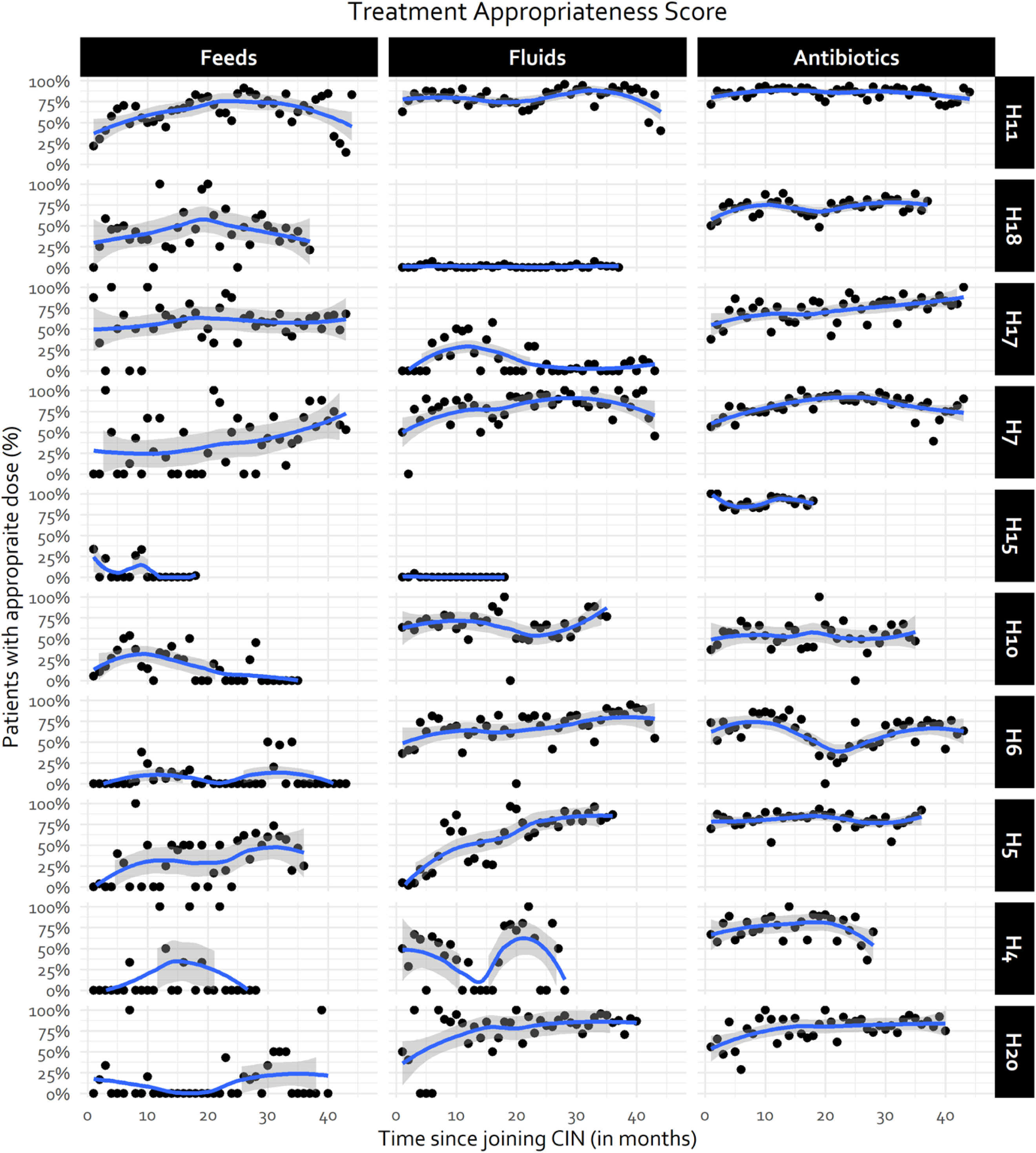
Hospital-specific treatment accuracy trends for half of randomly selected CIN-N hospitals. Trend line generated using LOWESS technique. Fewer observations in some hospitals due to different CIN-N joining dates.

**Supplementary Figure 3:**
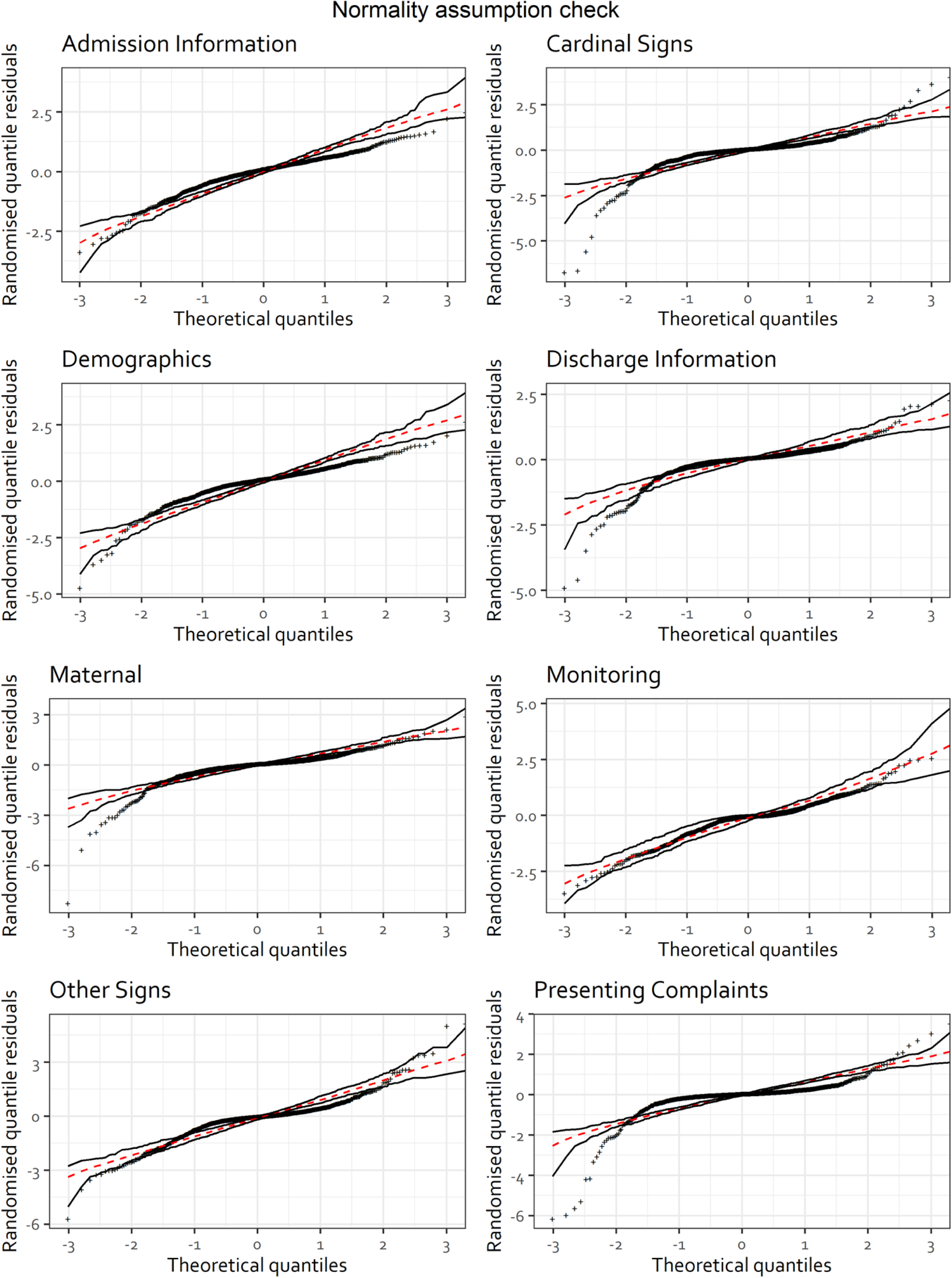
Normality assumption check for Poisson and negative binomial generalised linear mixed models for documentation completeness domains

**Supplementary Figure 4:**
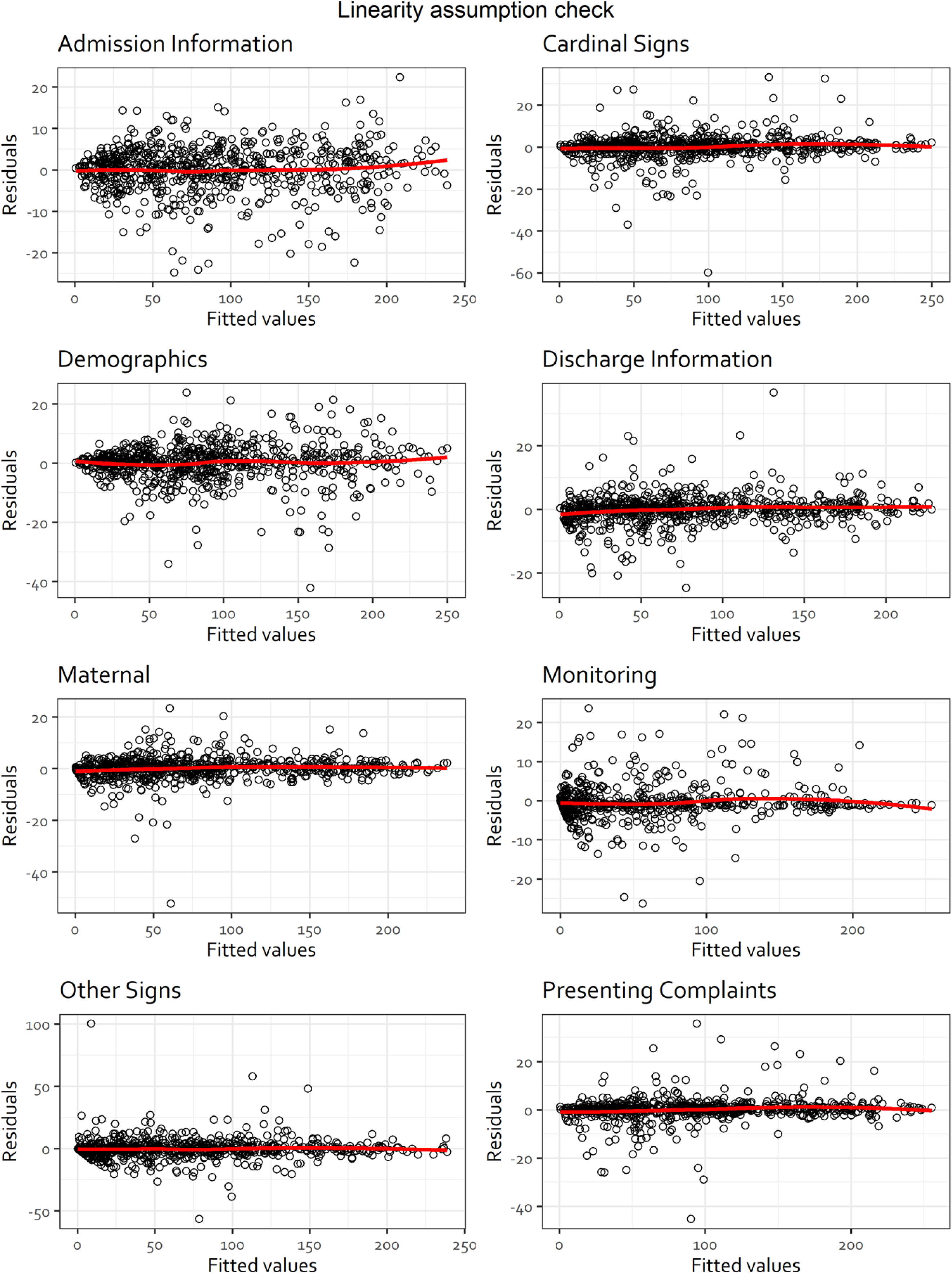
Linearity assumption check for Poisson and negative binomial generalised linear mixed models the documentation completeness domains

**Supplementary Figure 5:**
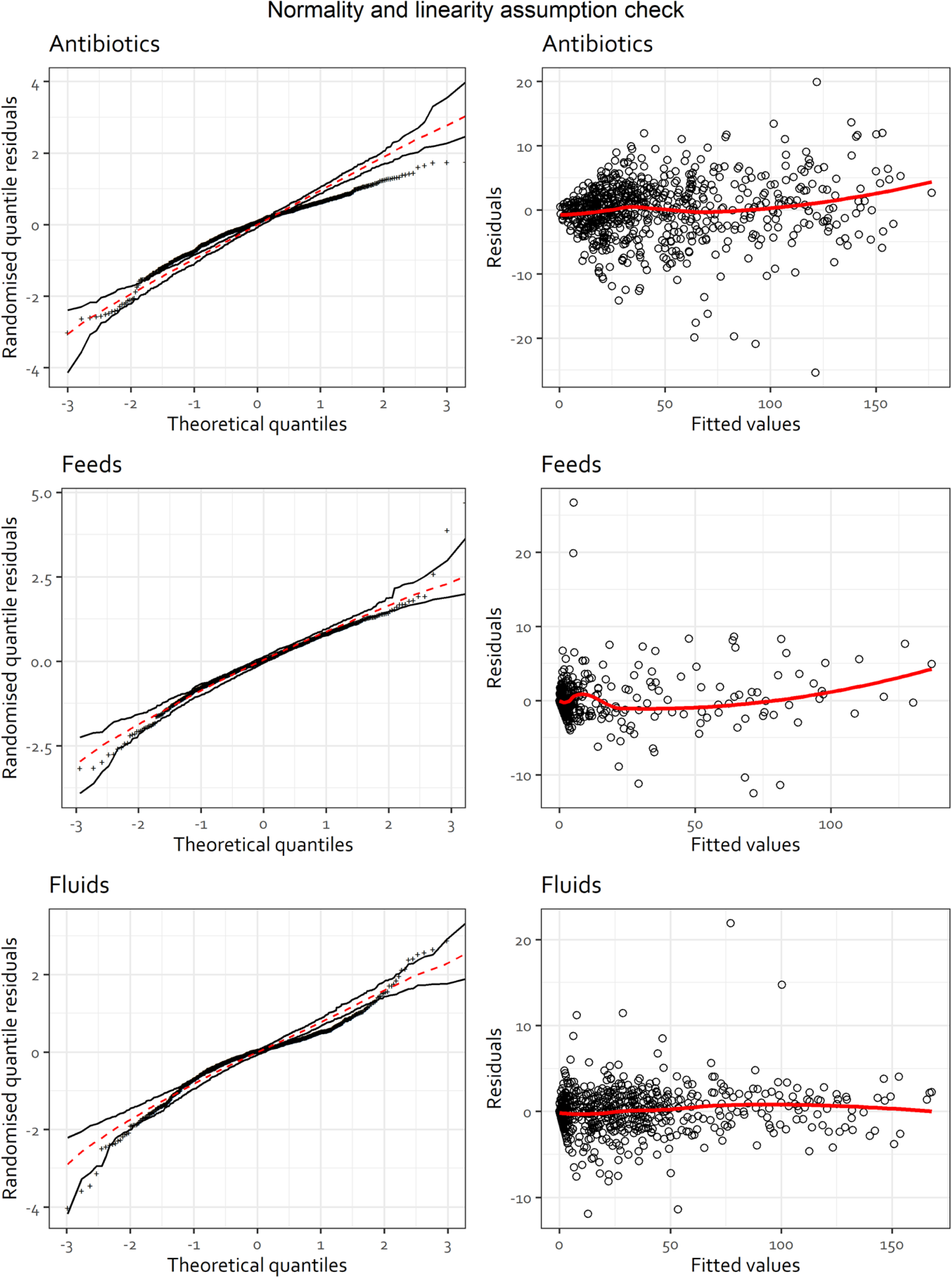
Linearity and normality assumption check for the treatment appropriateness Poisson and negative binomial generalised linear mixed models

